# Implementation, adoption and perceptions of telemental health during the COVID-19 pandemic: a systematic review

**DOI:** 10.1101/2021.07.05.21260018

**Authors:** Rebecca Appleton, Julie Williams, Norha Vera San Juan, Justin J. Needle, Merle Schlief, Harriet Jordan, Luke Sheridan Rains, Lucy Goulding, Monika Badhan, Emily Roxburgh, Phoebe Barnett, Spyros Spyridonidis, Magdalena Tomaskova, Jiping Mo, Jasmine Harju-Seppänen, Zoë Haime, Cecilia Casetta, Alexandra Papamichail, Brynmor Lloyd-Evans, Alan Simpson, Nick Sevdalis, Fiona Gaughran, Sonia Johnson

**Affiliations:** Mental Health Policy Research Unit, Division of Psychiatry, UCL; Centre for Implementation Science, King’s College London; Mental Health Policy Research Unit, King’s College London; Centre for Health Services Research, City, University of London, London, United Kingdom; Camden and Islington NHS Foundation Trust; Kingston iCope, Camden & Islington NHS trust; Division of Psychiatry, UCL; Centre for Outcomes Research and Effectiveness, Research Department of Clinical, Educational and Health Psychology, University College London, London, UK; South London and Maudsley NHS Foundation Trust; Institute of Psychiatry, Psychology and Neuroscience, King’s College London; King’s Improvement Science, King’s College London

**Keywords:** telemental health, COVID-19, remote care, telemedicine, mental health, systematic review, implementation science

## Abstract

**Background:** Early in 2020, mental health services had to rapidly shift from face-to-face models of care to delivering the majority of treatments remotely (by video or phone call or occasionally messaging) due to the COVID-19 pandemic. This resulted in several challenges for staff and patients, but also in benefits such as convenience or increased access for people with impaired mobility or in rural areas. There is a need to understand the extent and impacts of telemental health implementation, and barriers and facilitators to its effective and acceptable use. This is relevant both to future emergency adoption of telemental health, and to debates on its future use in routine mental health care.

**Objective:** To investigate the adoption and impacts of telemental health approaches during the COVID-19 Pandemic, and facilitators and barriers to optimal implementation.

**Methods:** Four databases (PubMed, PsycINFO, CINAHL and Web of Science) were searched for primary research relating to remote working, mental health care, and the COVID-19 pandemic. Preprint servers were also searched. Results of studies were synthesised using framework synthesis.

**Results:** A total of 77 papers met our inclusion criteria. In most studies, the majority of contacts could be transferred to a remote form during the pandemic, and good acceptability to service users and clinicians tended to be reported, at least where the alternative to remote contacts was interrupting care. However, a range of impediments to dealing optimal care by this means were also identified.

**Conclusions:** Implementation of telemental health allowed some continuing support to the majority of service users during the COVID-19 pandemic and has value in an emergency situation. However, not all service users can be reached by this means, and better evidence is now needed on long-term impacts on therapeutic relationships and quality of care, and on impacts on groups at risk of digital exclusion and how to mitigate these.

## Introduction

Since the onset of the COVID-19 pandemic in the first few months of 2020, most countries have experienced a severe disruption of mental health service delivery in its usual forms (1). Community-based outpatient, day and home treatment programmes, prevention and mental health promotion programmes, and services for specific age groups, such as older adults, children and adolescents and people with substance misuse problems, have been among those severely affected at a time of potentially increased demand due to the adverse mental health consequences of the pandemic (2, 3).

Mental health care providers around the world responded to the disruption of services in many ways, including the significant and widely documented shift to remote delivery of mental health services to replace in-person consultations (1, 4, 5). Telemental health, defined as “the provision of behavioral and/or mental health care services using technological modalities in lieu of, or in addition to, traditional face-to-face methods” (6), including video conferencing, telephone, email or text messaging, has been central to continuing assessment and support in the community. Additionally, technological innovations are helping to address isolation and service disruption in hospital and residential settings (4, 7).

Multiple research studies conducted both before and during the pandemic have reported evidence of the effectiveness of telemental health in reducing treatment gaps and improving access to care for a range of service users (8-10). Findings from studies, often of telemental health programmes established for purposes of research, have suggested that, overall, synchronous modalities such as video conferencing are comparable to face-to-face delivery in terms of quality of care, reliability of clinical assessments and treatment outcomes and adherence (11-15). Good levels of service user acceptance and satisfaction with telemental health services have also been reported (10). Successful adoption of telemental health has been described across a wide range of populations (adult, child and adolescent, older people, ethnic minorities), settings (hospital, primary care, community) and conditions (11, 13, 16). For certain populations, including some with autism and severe anxiety disorders, and those with physical disabilities or geographical barriers to accessing services, telemental health can be preferable for some service users (6, 17), although individuals experiencing significant social disadvantage or severe mental health problems, such as psychosis, have been found to benefit less (18). Research suggests that telemental health can also work for group interventions (19). The attitudes of clinicians who have delivered care via synchronous telemental health appear to be largely positive, with professionals finding it both effective and acceptable (20) and recognising its potential to enhance communication within and between mental health teams (4, 7). There is also some positive health economic evidence, with several studies suggesting telemental health is no more expensive than face-to-face delivery and tends to be more cost-effective (12). This approach also appears to be a viable and inexpensive treatment option where access to emergency services is limited, and associations have been found with reduced psychiatric admissions (10).

However, despite this evidence base, integration of telemental health approaches into routine mental health care or the widespread adoption of remote working across whole systems has rarely been reported. Even during the pandemic, adoption of such technologies has been piecemeal, with utilisation varying substantially both between and within countries (1, 7). Barriers to the wider adoption of telemental health include: the risk of digital exclusion of some service users, such as those facing significant social disadvantage or with limited technological access and expertise; lack of technological infrastructure and clear protocols within services, impeding the integration of telemental health with face-to-face care; difficulty in establishing and maintaining therapeutic relationships and in conducting high quality assessments; service users who lack private space or find participating in sometimes intimate and distressing discussions from home intrusive (4, 11, 12, 18, 21-23). A range of other ethical, regulatory, technological, cultural and organisational barriers have also been identified, both before and during the pandemic (12, 24-28).

The widespread emergency adoption of telemental health since the onset of the pandemic has generated a substantial literature. Numerous commentaries, service evaluations and reports of telemental health innovations and service user, carer and staff experiences, and a growing number of research studies addressing effectiveness and implementation issues (29-32), have been published internationally. Clinical guidelines have been rapidly produced in a number of countries (33).

A synthesis of the relevant empirical evidence gathered during the pandemic is therefore timely, and informative for future planning by generating evidence of effects of adopting telemental health across whole populations and service systems rather than in the context of relatively small-scale research studies involving volunteer participants. Capturing the learning and experiences gained through the rapid shift to telemental health will help optimise remote healthcare in a population that presents unique relational challenges associated with mental distress. It will also help to understand and overcome implementation barriers and inform strategies for improving the flexibility, effectiveness, and efficiency of mental health services through the sustained integration into routine care of telemental health approaches, to ensure that it brings the greatest benefits for patients, carers, and staff.

The aim of this rapid review is to synthesise the international literature specific to remote working in mental health services (as a replacement for or in conjunction with face-to-face service delivery) in the context of the COVID-19 pandemic. Our research questions are:

1. What evidence has been obtained during the COVID-19 pandemic regarding the effectiveness and cost-effectiveness of telemental health, and regarding its safety (including adverse events due to breaches of privacy and safety)?
2. What coverage has been achieved through telemental health adoption in the pandemic (including extent of adoption by services and reach among clinical populations in which it is adopted); in which groups and for which service settings is telemental health more or less likely to be implemented successfully; what are potential risks associated with digital exclusion for those not reached; and what barriers and facilitators influence success in implementation?
3. How acceptable are telemental health approaches to service users, carers and staff as applied during the pandemic, including perceived impacts on therapeutic relationships, communication and privacy?
4. What innovations and improvements have been introduced to make clinical care via telemental health more effective and acceptable, achieve greater coverage, and address barriers to delivering care in this way? (This includes descriptions and evaluations of specific strategies designed to make telemental health work better than usual delivery, and of adaptations of telemental health to specific settings, such as inpatient wards and crisis services).

## Methods

A systematic review was conducted following the Preferred Reporting Items for Systematic Reviews and Meta-Analyses (PRISMA) statement (34).

The protocol for this review was registered with PROSPERO (CRD42021211025).

### Search strategy

Four electronic databases (PubMed, PsycINFO, CINAHL and Web of Science), preprint servers medRxiv, PsyArXiv, Wellcome Open Research, and JMIR were searched for research relating to COVID-19, mental illness, and remote working from 1st January – 9th December 2020. An example search strategy can be found in Supplementary Table 1.

This search was supplemented by searching the references listed in included studies for any additional studies that met our inclusion criteria.

### Screening

The resulting list of articles was de-duplicated using Endnote (35) and all references were imported into Rayyan (36) for title and abstract screening. Full texts were sourced for articles deemed relevant for inclusion, and these were screened against the full review eligibility criteria. To establish consistency in study selection, title and abstract screening was conducted by four reviewers (MS, ZH, JHS & LSR), with 100% of included and 25% of excluded references checked by another reviewer (RA). Full texts were screened by three reviewers (RA, MB & MS), with 100% of included and 25% of excluded papers checked by another reviewer (task divided between LG, HJ, JW, PB & LSR). Any disagreements were resolved through team discussion.

### Inclusion criteria

#### Participants

Staff working within the field of mental health, people receiving organised mental health care for any condition (including addictions, dementia and intellectual disability), family members or carers of people receiving mental health care (regarding their views on the impact of remote working on the service user, and interventions aimed at reducing carer distress).

#### Interventions

Any form of spoken or written communication carried out between mental health professionals or between mental health professionals and service users / family members / unpaid carers or peer support communications using the internet, the telephone, text messaging platforms or hybrid approaches combining different platforms.

#### Comparator(s)/control

Any mental health communication delivered face-to-face, digitally or remotely, waitlist control or placebo. Studies comparing different modes of delivery during the pandemic, and those comparing care delivery and outcomes during the pandemic with those before the pandemic were included. Relevant studies with no comparator were also included.

#### Outcomes

Qualitative and quantitative outcomes describing: implementation effectiveness (including process evaluations) and barriers and facilitators to digital engagement, clinical effectiveness, cost-effectiveness, acceptability (including service user, carer and staff satisfaction), impacts on communication and therapeutic relationships, coverage and impacts of digital exclusion, interventions to improve quality or coverage, improvements in quality of life, and economic impacts.

#### Design

Any papers that present qualitative or quantitative data from study designs of any type (including relevant service evaluations and case series). If the focus of the study was not solely remote working but the results section contained substantial data relevant to our research questions, these were also included. Any relevant reviews identified in the searches were checked for included research which met our inclusion criteria.

### Exclusion criteria

We excluded studies that were:

a. not specific to the pandemic response.
b. reporting on interventions with patients with primary sleep disorders.
c. reporting on those with sub-clinical symptoms (unless combined with another included mental health problem).
d. focused on digital interventions such as apps, websites, and virtual reality tools, except where the sole purpose of the digital intervention was to facilitate direct spoken or written communication.
e. focused on interventions aimed at improving the mental health or wellbeing of healthcare professionals.
f. editorials, opinion pieces, guidance documents, protocols, conference abstracts and letters, with the exception of editorials or letters which contained primary research findings.

No language or location restrictions were applied in this review.

### Data extraction

Data extraction was supported by well-established implementation science frameworks. A data extraction form was developed based on a brief version of the Consolidated Framework for Implementation Research (CFIR) (37) and the taxonomy of implementation outcomes (38). We used the higher level CFIR constructs (see Table 2 for a brief definition of each one of the implementation facets that CFIR constructs capture) to extract data on factors influencing implementation success (39), and the taxonomy of implementation outcomes including acceptability, adoption and feasibility. We also extracted information deemed relevant based on previous studies conducted by the research team, including an umbrella review of pre-COVID 19 systematic reviews on telemental health and a qualitative study (15, 21). Data extracted consisted of study details, including design and focus of study; gender, ethnicity, age; diagnosis of participants; details of staff occupation; setting and context of study; intervention details, implementation outcomes (including acceptability, adoption, appropriateness, feasibility, fidelity, cost effectiveness, penetration and sustainability), barriers and facilitators to implementation and clinical and safety outcomes. The full data extraction form can be viewed in Supplementary File 1.

**Table 1.**
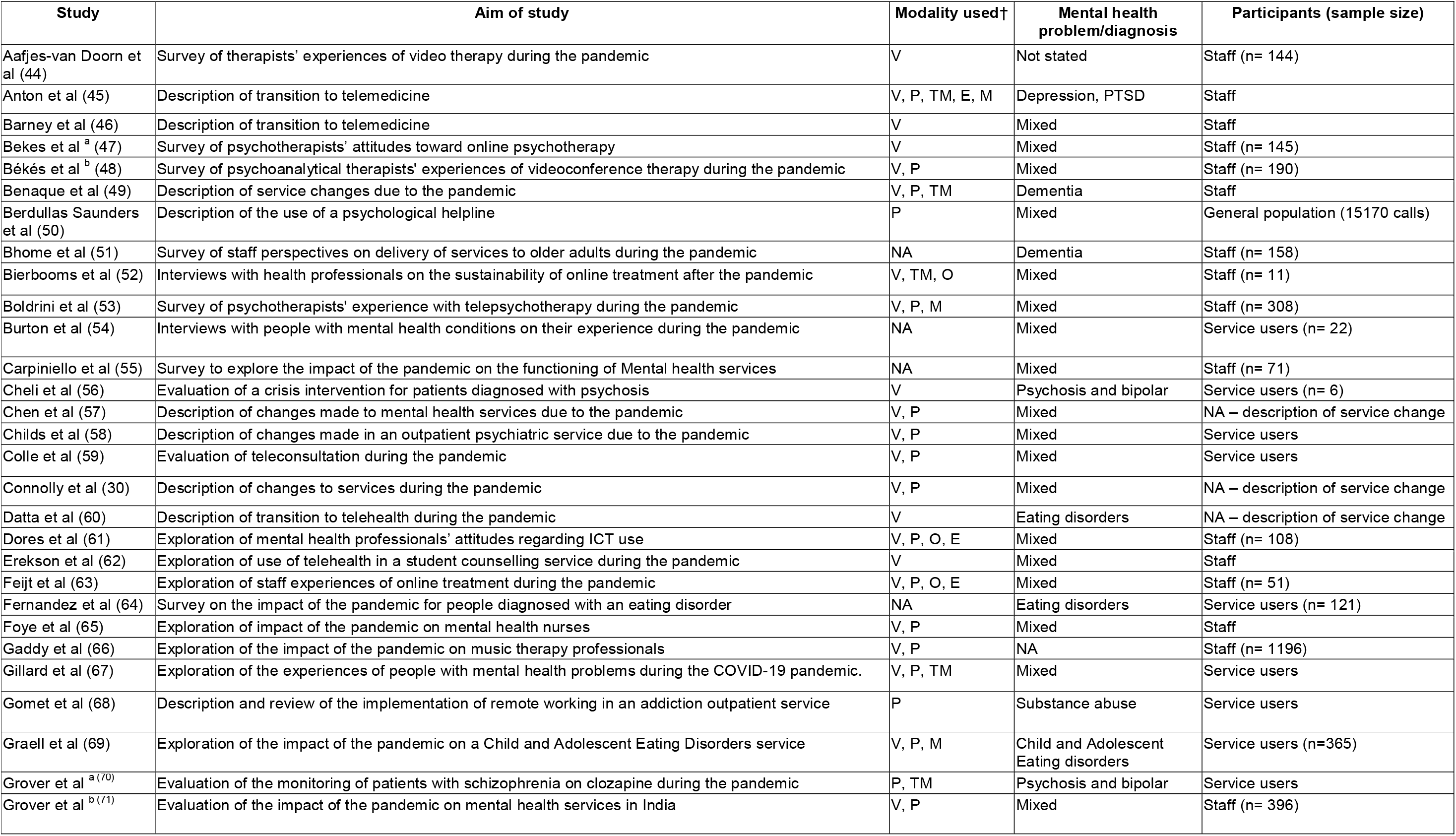
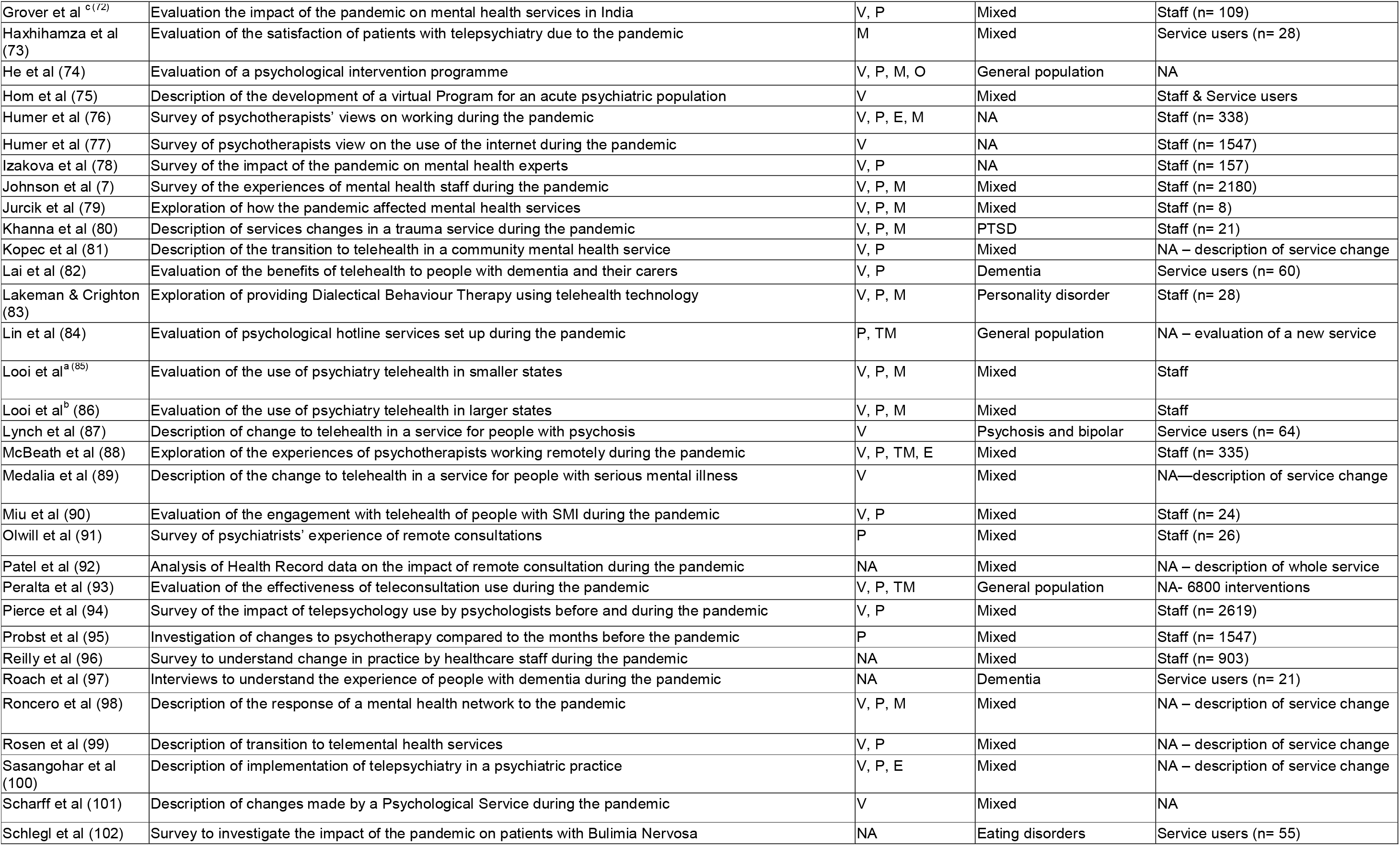
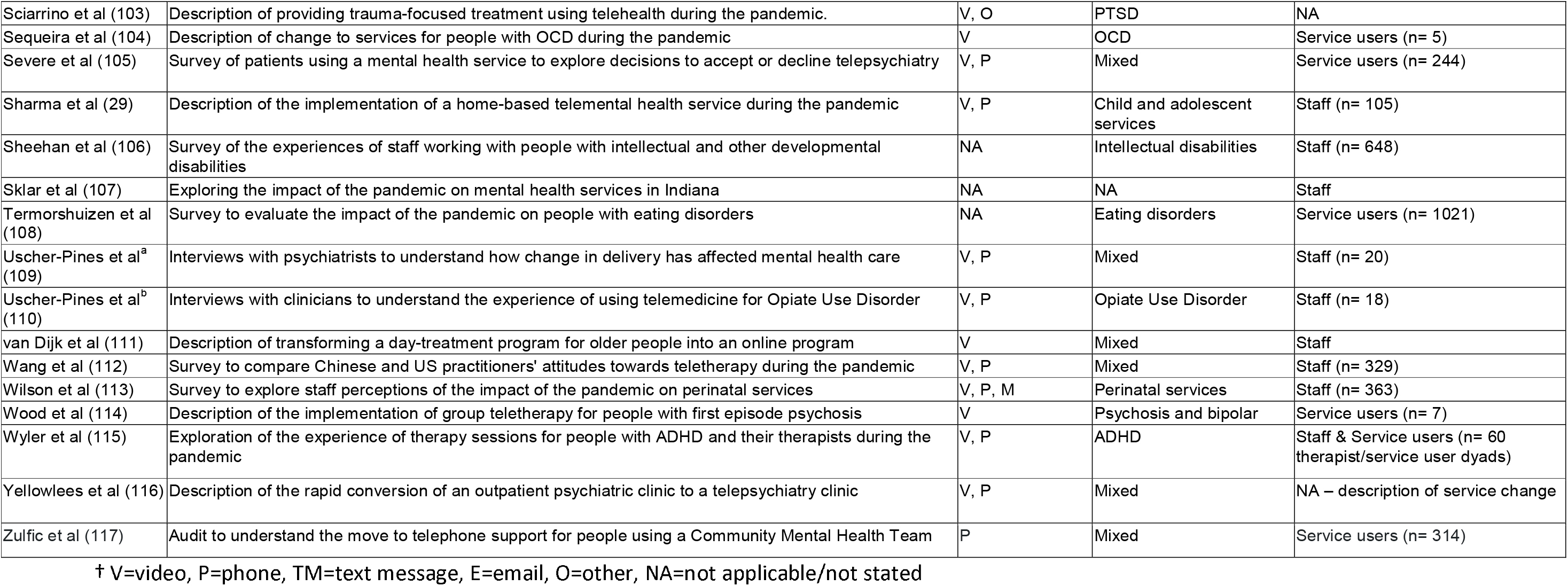
Study Characteristics

**Table 2.** Implementation barriers and drivers for telemental health grouped according to condensed CFIR domains (Consolidated Framework for Implementation Research)

| CFIR Domain | Findings | Example References |
| --- | --- | --- |
| Intervention Characteristics:<br>*Whether the intervention was internally/externally developed, evidence supporting the intervention, advantages compared to other methods of delivery, adaptability, trialability, and complexity* | - Remote care had advantages over face-to-face, e.g. making therapy more accessible for certain groups such as service users in remote locations; saving users money on travel; helping therapists get a better idea about the service users' home environment; some users benefitted from the distance, found it easier to communicate openly and became more independent.<br>- The main barriers for clinicians to deliver quality therapy were: picking up on non-verbal cues; assessing mental health symptoms; keeping service users engaged.<br>- Video and phone calls were the most common modalities; however, studies also reported the use of emails, instant messaging services, apps, videos and forums.<br>- Duration of telemental health appointments were shorter than face-to-face; clinicians reported it required more concentration and was more tiring.<br>- In some cases, studies have reported using shorter but more frequent appointments to deal with challenges in remote working (e.g. some service users struggling to stay focused). This was also used as a method to increase flexibility.<br>- Frequent contacts between sessions helped to build the therapeutic relationship. | (44, 52, 57, 87, 109, 110) |
| Outer Setting:<br>*Information on whether the organisation is networked with others, peer pressure to* | - Implementation was commonly due to 'stay at home' orders or national lockdowns, or a high level of COVID-19 cases in that area resulting in social distancing requirements.<br>- In the US, health insurers did not always cover telemental | (30, 57, 58, 76) |
| *implement intervention and external policies and incentives* | health care, whilst in some European countries, insurance cover for telemental health terminated at the end of the first wave of infections.<br>- Telehealth service delivery was eased by the relaxation of policy and billing reimbursements during this time.<br>- Professional bodies facilitated transition to telehealth by posting guidelines on their websites to assist clinicians.<br>- Platform developers worked rapidly to increase capacity.<br>- Clinicians identified the need for a video tool that adheres to privacy standards and links with a technical helpdesk.<br>- There were also concerns over the reduction in services to support the physical health needs of mental health service users. |  |
| *Inner Setting*<br>*Information on the structural characteristics, networks and culture of an organisation, as well as the implementation climate (e.g. capacity for change)* | - Overall, all settings had sufficient capacity to shift to some delivery of telemental health in a short period of time. | (77, 101, 103, 106) |
| *Staff Characteristics*<br>*Information on the following psychological attributes and also on any effects of staff demographic and professional backgrounds* | - There was some variation in acceptability of remote ways of working for staff depending on their therapeutic approaches.<br>- Telemental health take-up was dependent on: perceived experience of patient (positive or negative), comfort with online platform, previous clinical experience.<br>- Some staff felt less confident about professional skills during online compared to in-person consultations, especially those with less clinical experience and those who perceived their patients disliked remote care. | (44, 53, 112) |
| *Process*<br>*Training provided and any processes put in place to support telemental health intervention, planning and feedback on progress of implementation* | - The transition to telemental health occurred usually over a short time period.<br>- Training staff to use platforms was mentioned frequently, as was phoning service users to let them know about the transition to telemental health and how care would be provided going forwards.<br>- Methods of staff training included courses; shadowing or observing senior colleagues; discussion within clinical teams; facility-level telehealth coordinators; clinical champions providing training; and webinars.<br>- Sources of information for staff: colleagues; government guidelines; prepared consent forms; posts on listservs; APA and other official guidelines.<br>- New workflows had to be developed to allow staff to access patient records remotely.<br>- Despite some training, staff reported lack of support and identified training needs across several studies regarding: | (7, 29, 45, 46, 51, 60, 62, 63, 75, 106, 113, 116) |
|  | how to use online platforms and meeting privacy regulations. |  |
| Service User Needs/Resources<br>*Statements demonstrating awareness of the needs and resources of those served by the organization e.g. barriers and facilitators and feedback* | - A commonly reported issue was access to technology, particularly amongst service users with diagnoses such as schizophrenia, service users with a lower SES, and older adults (one study mentioned that older adults often lacked access to video software, so preferred phone calls).<br>- Concerns around privacy and confidentiality, and forming a therapeutic relationship may be more difficult when using remote care.<br>- Difficulties for service users to concentrate within a digital environment.<br>- Several studies mention the need for an agreed 'Zoom etiquette' for service users, including attire, audio/visual set-up and reducing background distractions.<br>- Stable internet connection was a problem for some service users.<br>- Some clients benefitted from the distance created by online treatment, as they became less inhibited and less dependent on therapist. | (51, 63, 78, 87, 99, 117) |

Data extraction was completed by nine reviewers (approximately eight studies each) using EPPI-Reviewer 4 (40). All reviewers were trained on how to extract data to ensure consistency, and extracted data was checked by a second reviewer. The extraction form was first piloted on 10% of included studies to assess usability and content, with amendments made before completing extraction for the whole dataset.

### Quality appraisal

Given the diversity of the included article types and methods, two quality appraisal tools were used. Primary research studies were assessed with the Mixed Methods Appraisal Tool (MMAT) (41). Commentaries and service evaluations were assessed using AACODS, which appraises the veracity, clarity, acknowledgement of bias, and relevance of the contribution to the field (42). Study quality was assessed by RA and verified by NVSJ. Disagreements were resolved through discussion.

### Evidence synthesis

We conducted a framework synthesis of study characteristics and outputs. Study outcomes were tabulated applying existing implementation science frameworks, i.e., the CFIR framework (37), Proctor et al.’s (38) taxonomy of implementation outcomes; and also relevant topics/themes that emerged during data extraction. This table-based synthesis of the study outcomes combined a deductive and inductive approach to data analysis by using existing frameworks, while at the same time, identifying emerging themes. Results reported in this paper include a narrative synthesis of the study characteristics and quantitative study outputs (43) and the tabulated results.

## Results

### Study selection

A total of 3956 references were identified through searching databases of published papers. medRxiv was the only preprint database where included papers were found (n=10), one more relevant paper was identified by a member of the research team and a further paper was found through reference searching of included studies.

A PRISMA flow diagram (34) of the screening and selection process is presented in Figure 1.

**Figure 1:**
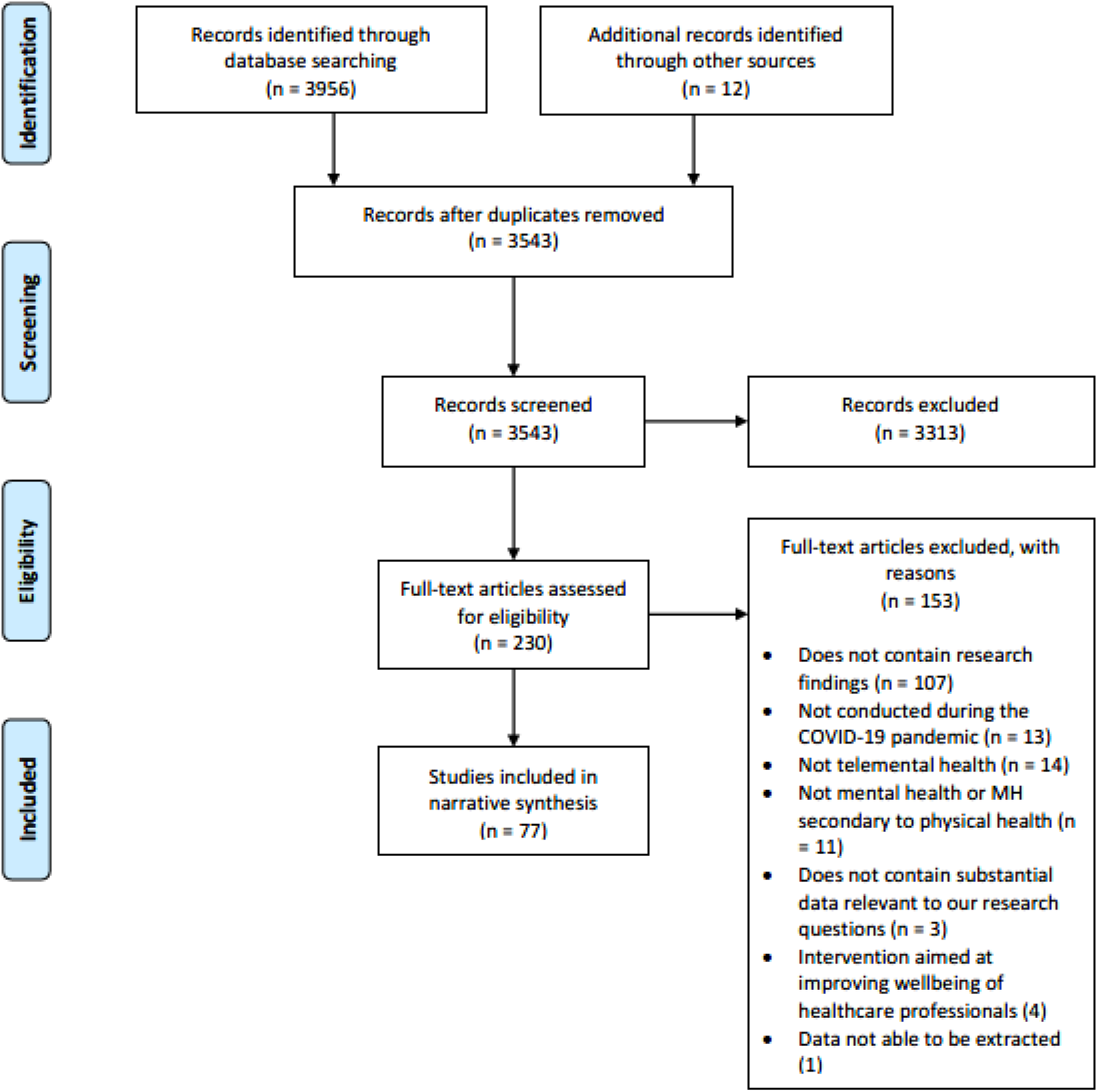
PRISMA diagram showing screening and included studies

### Quality of included studies

The quality of included primary research studies was moderate to high: 23 out of 48 studies appraised using the MMAT met above 80% of quality criteria, whilst 21 out of 48 met between 50 and 79%. The quality of included service evaluations or audits were generally high: 27 out of 29 studies appraised using the AACODS met at least four out of the six quality criteria assessed. These include: being written by recognised experts, including reference lists, having a clear aim, stating details such as date, location and limitations, and making a meaningful contribution to the research literature.

### Study characteristics

Of the 77 studies which were eligible for inclusion in the review, 45 were primary research studies, 24 were service evaluations or audits and eight were editorials or letters that included data. Thirty-three were conducted in the USA, nine in the UK, and five each in Australia, Canada and Spain. Five were conducted across more than one country.

Of the 45 primary research studies, 32 involved staff and 9 involved service users. The remaining four analysed service use data (three evaluated contacts with hotlines and one evaluated service use in one UK National Health Service (NHS) service provider).

Most studies were conducted in services that worked with people with mixed psychiatric diagnoses (n=30), although we also found studies conducted with groups with a single diagnosis, e.g. dementia or eating disorders. Studies could include more than one service type, the most commonly studied being community mental health teams (CMHTs) or outpatient settings (n=39), followed by psychology or psychotherapy services (n=17). Inpatient or residential services were included in 15 studies, whilst 12 included general hospitals. Eight studies included private hospitals or clinics, whilst four explored telemental health use in helplines, voluntary sector services, crisis teams, or veterans’ health services respectively. Five studies did not report any specific setting.

The aims of most studies were either a description of changes made due to the pandemic, new services set up because of the pandemic, or an evaluation of the impact of the pandemic on either staff or service users. The descriptions of changes either focused specifically on the move to telemental health or were wider descriptions of changes to services including the use of telemental health. The characteristics of each of the included studies are shown in Table 1, with a more detailed summary in Supplementary Table 2.

### Data synthesis

#### Barriers and facilitators to telemental health

Implementation barriers and facilitators were categorised using a condensed version of the CFIR framework (see Table 2, where definitions of the CFIR constructs are also provided). The key findings are summarised below.

#### Intervention characteristics

Video and phone calls were the most common modalities used for remote care; studies also reported the use of emails, instant messaging services, apps, pre-recorded videos and forums (further details about the modality used in each study can be found in Table 1).

When comparing remote care to traditional face-to-face settings, studies identified advantages for both methods. Benefits for remote care included being more convenient (for both staff and service users), making care more accessible to groups who may previously have been excluded, reducing travel (resulting in both time and cost savings), and helping clinicians understand more about the service user, as they had more insight into their home lives. A further benefit is that more family members were readily able to attend family therapy or family education sessions since care was moved online e.g. (44). However, clinicians reported difficulties in picking up on non-verbal cues in remote compared to face-to-face care, and that remote care could sometimes require more concentration.

#### Outer Setting

Services commonly implemented remote methods of working due to ‘stay at home’ orders or national lockdowns, or due to a high level of cases in their local area. Overall, all settings described in papers had sufficient capacity to make a rapid shift to remote forms of care. Several studies in the USA in particular mentioned the impact of health insurance regarding uptake of telemental health e.g. (57), as not all insurance providers covered remote care. However, this did change during the course of the pandemic as telemental health delivery was eased by the relaxation of policy and billing reimbursements (57, 76). The change from face-to-face to remote delivery of care was also facilitated by professional societies who posted guidelines on their websites to assist clinicians.

#### Staff Characteristics

Enablers for clinician uptake included supporting clinicians by ensuring supervision, supportive leadership, clear communication, keeping track of clinicians’ needs, optimising physical space for comfort and privacy (for example using headphones or ergonomic seating), and arranging times away from the computer.

However, staff in several studies reported a lack of initial training for telemental health, and therefore identified training needs regarding the use of online platforms and meeting privacy regulations in particular. In some studies, having no previous experience with telemental health was also found to be associated with higher anxiety (44) and lower uptake (53) of remote care. However, others found previous experience did not impact clinicians’ views of telemental health during the pandemic (48).

#### Process

As telemental health was not commonly used in most services pre-COVID, staff had to rapidly adjust to a new way of working. Several studies discussed the training which was put in place for staff, which included training courses, shadowing or observing senior colleagues, discussion within clinical teams, facility-level telehealth coordinators and clinical champions providing training, webinars, and checking official guidelines. New workflows also had to be developed to allow staff to access patient records remotely, and service users had to be informed about the transition to telemental health.

#### Service user needs/resources

In addition to the needs of staff, service users also identified certain needs and resources to enable them to effectively transition to telemental health care. A commonly reported issue was access to technology, particularly amongst service users with diagnoses such as schizophrenia e.g. (117), older adults e.g. (51), and service users from lower socio-economic backgrounds e.g. (79). Service users also reported problems having a stable internet connection to allow for uninterrupted communication, which could negatively impact the therapeutic relationship. Concerns were also raised by both clinicians and service users regarding privacy and confidentiality, and in some cases service users had difficulties concentrating on remote care. Several studies e.g. (87, 99), mention the need for an agreed ‘Zoom etiquette’ for service users, including attire, audio/visual set-up and background distractions.

### Implementation outcomes

Outcomes of the implementation of telemental health have been summarised below using Proctor et al.’s (38) taxonomy of implementation outcomes. Further information can be found in Table 5.

#### Acceptability

Remote care was seen as satisfactory by the majority of clinicians and service users in most studies in the context of the pandemic. A number of studies also reported that telemental health enabled some groups to access care who found it difficult to engage with face-to-face support e.g. (7). Some clinicians reported that they would also be willing to continue with some aspects of remote care in the future e.g. (44, 78). However, it is important to note that whilst acceptability was high overall, this was not the case for all groups e.g. Grover et al (72) reported acceptability rates of around 45% for both clinicians and service users using services in a range of settings in India. Further details of satisfaction and acceptability outcomes are shown in Table 3.

**Table 3:** Levels of Acceptability of telemental health during the COVID-19 pandemic

| Author | Service location and type | Acceptability data |
| --- | --- | --- |
| Aafjes-van Doorn et al | **Type of service:** Psychology/psychotherapy/counselling service<br>**Country:** USA, Canada Europe: Hungary, Italy, UK, Germany, Norway, Sweden, Switzerland, Latvia, Ireland | **Clinician views**<br>- *Mainly positive attitudes towards video therapy were reported ($M = 3.42$, $SD = 0.50$, range: 2.31–4.69). Views on video therapy had become more positive since the pandemic ($t(140) = 2.06$, $p < .05$); video therapy was still viewed as somewhat less effective compared to in-person therapy ($M = 2.19$, $SD = 0.65$, range: 1.00–4.00).*<br>**Service user and carer views (reported by clinicians)**<br>- *Only 7% ($n=10$) thought their patients experienced video therapy negatively ($N = 10$; 7.1%). The majority perceived patient experience as either positive ($N = 88$; 63.8%) or neutral ($N = 40$; 28.4%).* |
| Bekes et al (a) | **Type of service:** Psychology/psychotherapy/counselling service; Private hospital/clinic; CMHT and outpatient services<br>**Country:** Canada, USA, Europe (countries not stated) | **Service user and carer views (reported by clinicians)**<br>- *Psychotherapists reported that their patients had an extremely positive ($N=20$; 13.8%), positive ($N=71$; 49%), or neutral ($N=40$; 27.6%) experience with online psychotherapy. 7.6% of the psychotherapists thought that their patients experienced online psychotherapy somewhat negatively and none of the psychotherapists reported an extremely negative patient experience.* |
| Békés et al (b) | **Type of service:** CMHT and outpatient services; Psychology/psychotherapy/counselling service; Private hospital/clinic<br>**Country:** Canada, USA, Europe (countries not stated) | **Clinician views**<br>- *Challenges included technical/internet problems (64.7%), patients not having a private space, (46.8%), risk of patient (44.7%) or therapist (26.3%) getting distracted, difficulty feeling connected to patients (29.5%) or reading their emotions (27.4%), difficulty keeping professional boundaries (23.2%) & confidentiality concerns (16.3%). 64.2% ($n = 122$) reported their relationships with service users felt as authentic to pre-COVID, 46% felt as emotionally connected & 64% reported no change to the therapeutic relationship.*<br>**Service user and carer views (reported by clinicians)**<br>- *Most therapists reported a positive ($n = 101$; 53.2%) or neutral ($n = 55$; 28.9%) patient experience, with only 34 reporting a somewhat negative online therapy experience for their patients (25.8%).* |
| Benaque et al | **Type of service:** Voluntary sector/non-profit<br>**Country:** Spain | **Clinician views**<br>- *81% of clinical staff considered the quality of telemedicine consultations to be either good or excellent. 75% viewed telemedicine visits as equal or better than face-to-face consultations.* |
| Colle et al | **Type of service:** CMHT and outpatient services<br>**Country:** France | **Clinician views**<br>- *94.1% of psychiatrists were satisfied with teleconsultations in this context.*<br>**Service user and carer views**<br>- *89.5% of patients were satisfied, 73.3% of patients spontaneously expressed their gratefulness for remote care.* |
| Dores et al | **Type of service:** | **Clinician views** |
|  | Psychology/psychotherapy/<br>counselling service<br>**Country:** Portugal | - 21 (out of 71) psychologists (29.6%) considered their experiences to be neither negative nor positive.<br>- Most of the respondents considered their experience with digital technologies to be either positive (n = 37; 52.1%) or very positive (n = 13; 18.3%). None reported their experiences as negative. |
| Grover et al<br>(a) | **Type of service:**<br>CMHT and outpatient services<br>**Country:** India | **Service user and carer views**<br>- 75.5% of patients and family members were satisfied they could remain in touch with the treating doctor.<br>- 1/4 of patients had difficulty in procuring clozapine, with clozapine not being available in their locality in 15% of cases & 3.4% having to switch their brand. 1/4 were able to get the absolute neutrophil count done in the previous month. |
| Grover et al<br>(b) | **Type of service:**<br>CMHT and outpatient services<br>Inpatient mental health service<br>Private hospital/clinic<br>**Country:** India | **Service user and carer views (as reported by clinicians)**<br>- 21% reported non-Healthcare workers (HCWs) in quarantine were dissatisfied, 19.9% reported that HCWs in quarantine were dissatisfied, and 13.5% reported that HCWs working with COVID patients were dissatisfied.<br>**Clinician views**<br>- Participants rated their satisfaction with the services they were currently providing to their patients with a mean of 45.8% on a Likert scale from 0-100 (SD = 28.6). |
| Grover et al<br>(c) | **Type of service:**<br>Medical colleges, government-funded<br>institutes; general hospital psychiatry<br>units **Country:** India | **Clinician views**<br>- Overall satisfaction with the mental health services being catered, the participants rated their level of satisfaction as 46.6% (standard deviation: 27.6). |
| Haxhihamza<br>et al | **Type of service:**<br>Day hospital<br>**Country:** Macedonia | **Service user and carer views**<br>- 20/28 strongly agreed/agreed that the medical care received was just about perfect. 4 patients agreed that they were dissatisfied with some things about their medical care (1 strongly agreed, 3 agreed).<br>- 20 (strongly) agreed that they can get medical care whenever they need it. 20 (strongly) agreed that they have easy access to medical specialists. 4 (strongly) agreed that the wait for emergency treatment was too long. |
| He et al | **Type of service:**<br>Helplines; Online media programs<br>**Country:** China | **Service user and carer views**<br>- Feedback from clients demonstrated that more than 50% felt their negative emotions, such as anxiety and depression, were relieved. |
| Hom et al | **Type of service:**<br>Private hospital/clinic<br>**Country:** USA | **Service user and carer views**<br>- Patients who have discharged thus far (n=10) have also expressed confidence in their aftercare plans. 2 patients who completed the exit survey reported very positive experiences & both rated their care as 9/10. |
| Izakova et<br>al | **Type of service:**<br>CMHT and outpatient services<br>Inpatient mental health service<br>**Country:** Slovakia | **Clinician views**<br>- 69.4% of them have considered it as an adequate form for diagnostics and therapy in the common clinical practice.<br>- 51.6% want to use it at a limited level with the defined guidelines also in future. |
| Johnson et | **Type of service:** | **Clinician views** |
| al | All service settings, including inpatient, CMHTs, voluntary sector.<br>**Country:** UK | - A majority (818, 74.0% of respondents) agreed/strongly agreed that video calls were suitable to assess progress of existing service users, but only 39.8% (442) agreed/strongly agreed that they were suitable for making the initial assessments. A majority (725, 65.8%) agreed/strongly agreed that use of remote care had resulted in not having contact with some service users who had not engaged with remote appointments. |
| Lakeman & Crighton | **Type of service:** Psychology/psychotherapy/ counselling service<br>**Country:** Australia | **Clinician views**<br>- 32% (n = 7) stated they were not confident at all in delivering online DBT, 50% (n = 11) reported being 'a little' confident and 4 reported feeling confident doing so. 14 respondents identified limited access to the internet, appropriate devices and/or internet blackspots as being significant obstacles to engagement. |
| Lynch et al | **Type of service:** CMHT and outpatient services<br>**Country:** USA | **Service user and carer views**<br>- The telehealth acceptance rates of the CP subsample indicated that 90% (n = 18) enrolled at the time of conversion agreed to telehealth sessions within ten days of the service transition. |
| Olwill et al | **Type of service:** CMHT and outpatient services<br>**Country:** Ireland | **Clinician views**<br>- 92% of respondents (n = 24) (and 100% consultants (n = 12)) reported lower confidence in making a diagnosis. 96%, (n = 25) agreed the lack of visual cues affected their assessment of the patient.<br>- 70% agreed that they found it more difficult to consider discharging a patient.<br>- 88% agreed they found it more difficult to establish a therapeutic alliance with new patients. |
| Sheehan et al | **Type of service:** CMHT and outpatient services<br>**Country:** UK | **Clinician views**<br>- 53.3% reported concerns of having to adapt too quickly to new ways of working; 37.9% reported having to learn new technologies too quickly and/or without sufficient training or support.<br>- 45.3% raised concerns around engaging patients with learning difficulties &/or autism. 23.7% had concerns around safeguarding or risk management. 27.9% reported greater workload than usual. |
| Wang et al | **Type of service:** Not stated/unclear<br>**Country:** USA & China | **Clinician views**<br>- Pre-COVID, 25% of US psychoanalytic practitioners felt mainly negative about teletherapy and 36% felt mainly positive, as compared to only about 9% and 47% of China American Psychoanalytic Alliance (CAPA) practitioners respectively.<br>- During the pandemic about 23% of US psychoanalytic practitioners felt mainly negative about teletherapy and about 37% felt mainly positive, compared to about 2% and about 58% of CAPA practitioners respectively. |
| Wilson et al | **Type of service:** CMHT and outpatient services; Crisis and emergency mental health services<br>Inpatient mental health service<br>**Country:** UK | **Clinician views**<br>- Staff reported feeling less able to assess women attending the perinatal mental health service using telemedicine, particularly their relationship with their baby (43.3%; 90/208), and to mobilise safeguarding procedures (29.4%; 62/211). |

Telemental health services were acceptable to people during the pandemic as a way of continuing their treatment, however findings from several studies also indicated that participants wanted at least some appointments to be face-to-face once restrictions on in-person contact had loosened.

#### Adoption

Adoption rates were relatively high across studies, with most services or clinicians moving their appointments to remote methods. Rates of adoption of telemental health for service users who were already receiving care at the start of the pandemic ranged from 48% (90) to 100% (45, 68, 111). Some studies reported face-to-face appointments still took place if necessary, for example for initial assessments or for medication reviews e.g. (117). Most studies which examined impact on attendance reported no adverse effects on attendance rates after introducing telemental health: there was either no difference in missed appointments when comparing remote to face-to-face care (46, 87), or non-attendance after adoption of telemental health decreased (80, 92, 104). Further details about adoption of telemental health across studies can be found in Table 4.

**Table 4:**
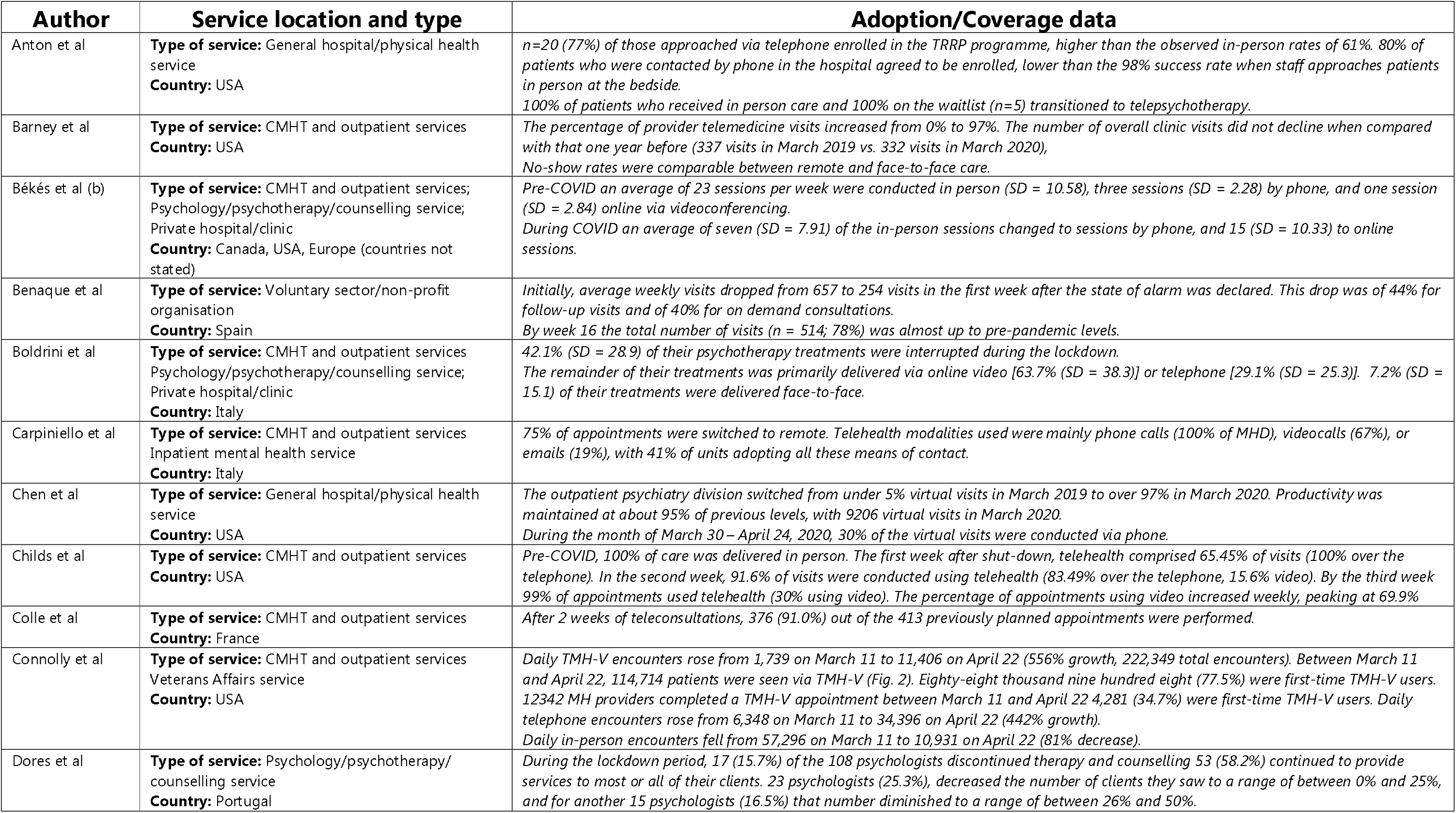
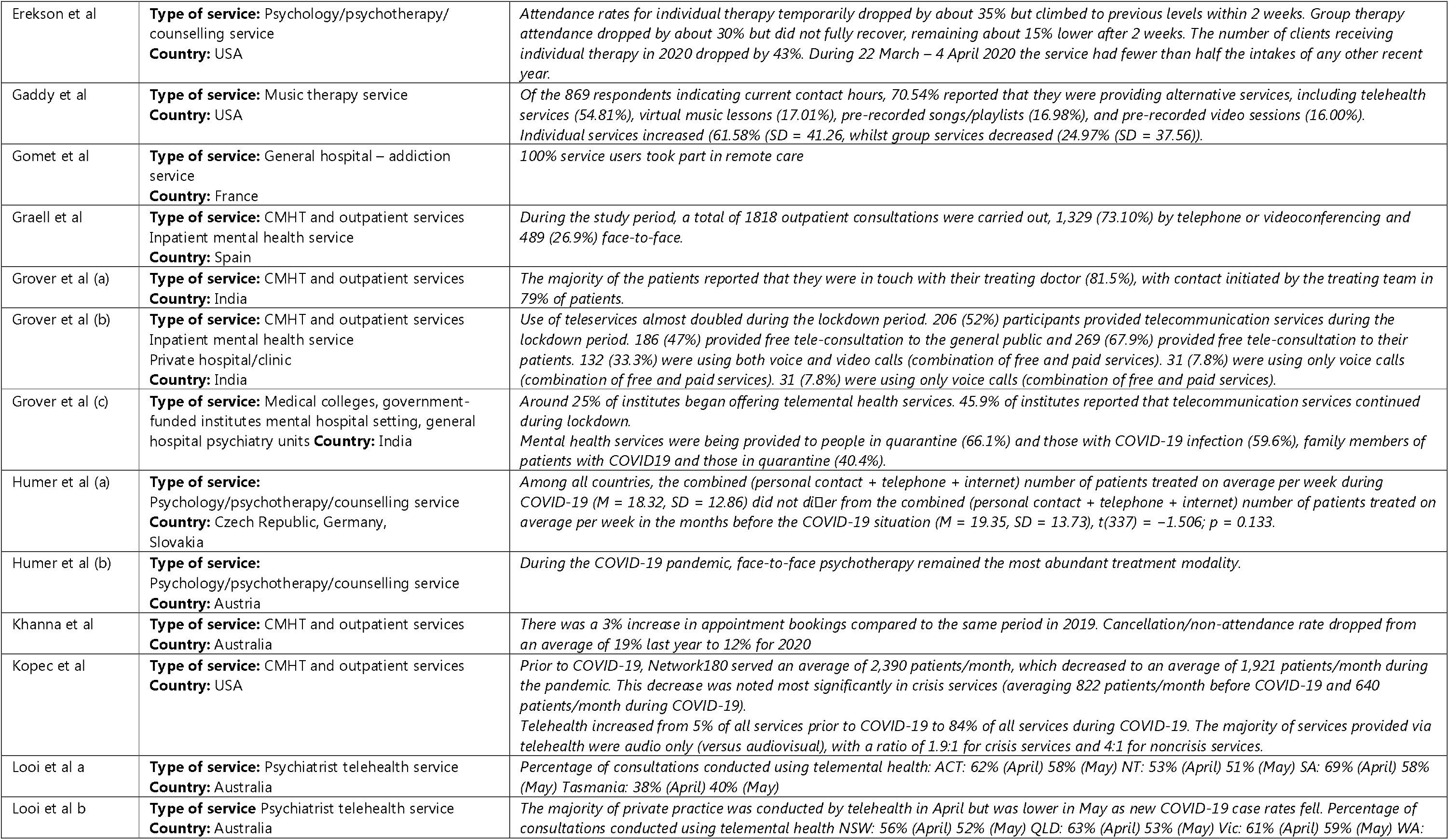
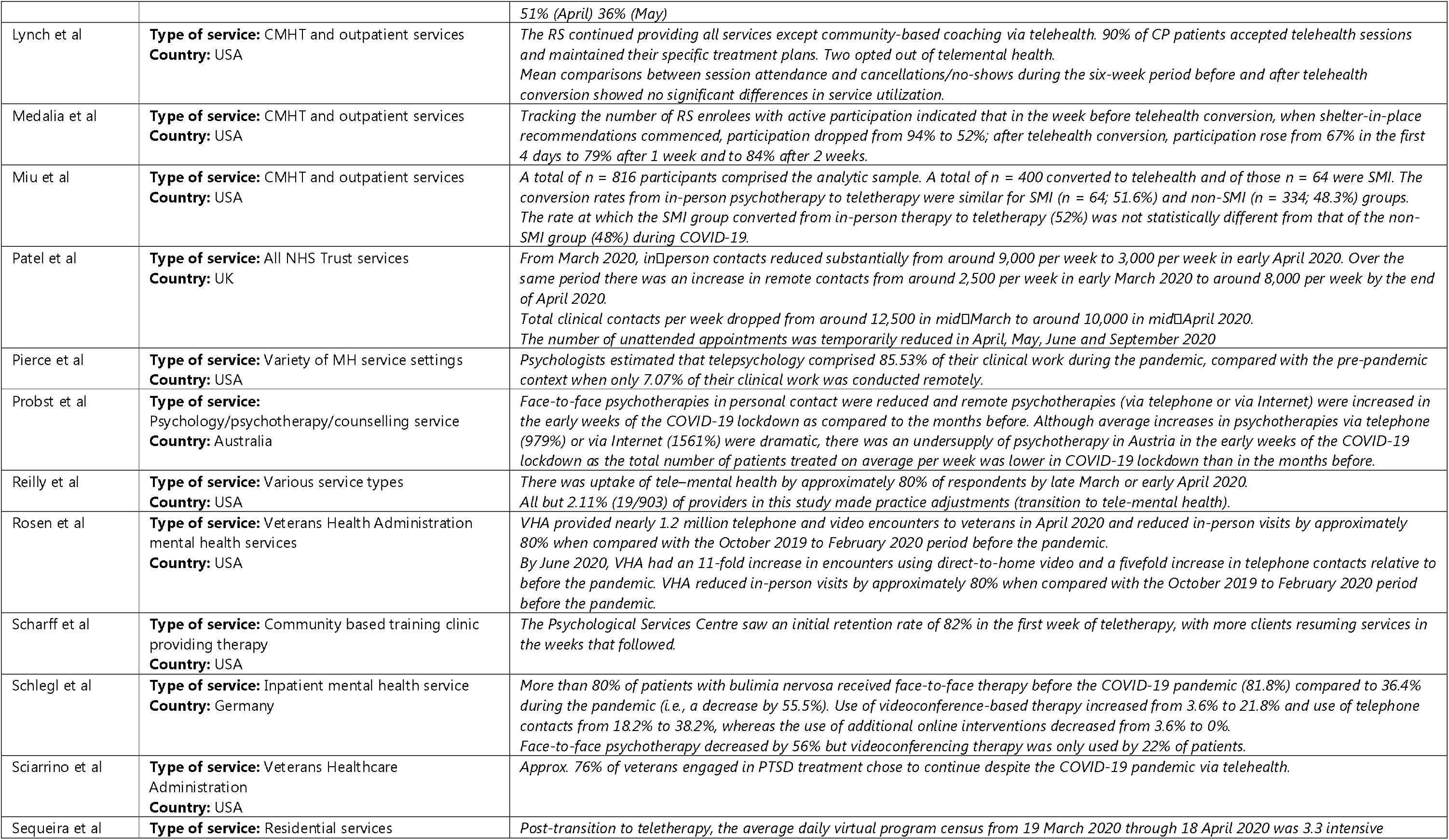
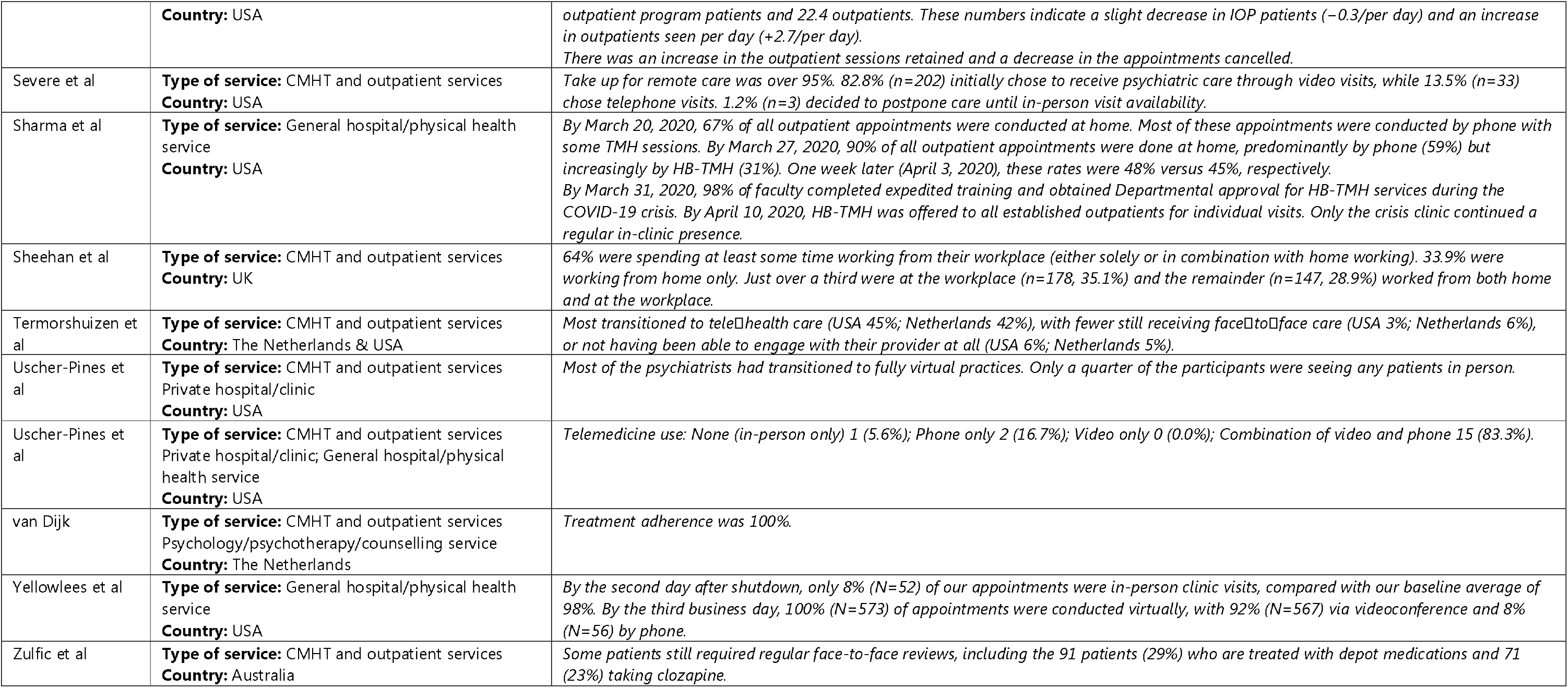
Levels of Adoption and Coverage of telemental health during the COVID-19 pandemic

| Author | Service location and type | Adoption/Coverage data |
| --- | --- | --- |
| Anton et al | **Type of service:** General hospital/physical health service<br>**Country:** USA | *n=20 (77%) of those approached via telephone enrolled in the TRRP programme, higher than the observed in-person rates of 61%. 80% of patients who were contacted by phone in the hospital agreed to be enrolled, lower than the 98% success rate when staff approaches patients in person at the bedside.*<br>*100% of patients who received in person care and 100% on the waitlist (n=5) transitioned to telepsychotherapy.* |
| Barney et al | **Type of service:** CMHT and outpatient services<br>**Country:** USA | *The percentage of provider telemedicine visits increased from 0% to 97%. The number of overall clinic visits did not decline when compared with that one year before (337 visits in March 2019 vs. 332 visits in March 2020),*<br>*No-show rates were comparable between remote and face-to-face care.* |
| Békés et al (b) | **Type of service:** CMHT and outpatient services;<br>Psychology/psychotherapy/counselling service;<br>Private hospital/clinic<br>**Country:** Canada, USA, Europe (countries not stated) | *Pre-COVID an average of 23 sessions per week were conducted in person (SD = 10.58), three sessions (SD = 2.28) by phone, and one session (SD = 2.84) online via videoconferencing.*<br>*During COVID an average of seven (SD = 7.91) of the in-person sessions changed to sessions by phone, and 15 (SD = 10.33) to online sessions.* |
| Benaque et al | **Type of service:** Voluntary sector/non-profit organisation<br>**Country:** Spain | *Initially, average weekly visits dropped from 657 to 254 visits in the first week after the state of alarm was declared. This drop was of 44% for follow-up visits and of 40% for on demand consultations.*<br>*By week 16 the total number of visits (n = 514; 78%) was almost up to pre-pandemic levels.* |
| Boldrini et al | **Type of service:** CMHT and outpatient services<br>Psychology/psychotherapy/counselling service;<br>Private hospital/clinic<br>**Country:** Italy | *42.1% (SD = 28.9) of their psychotherapy treatments were interrupted during the lockdown.*<br>*The remainder of their treatments was primarily delivered via online video [63.7% (SD = 38.3)] or telephone [29.1% (SD = 25.3)]. 7.2% (SD = 15.1) of their treatments were delivered face-to-face.* |
| Carpiniello et al | **Type of service:** CMHT and outpatient services<br>Inpatient mental health service<br>**Country:** Italy | *75% of appointments were switched to remote. Telehealth modalities used were mainly phone calls (100% of MHD), videocalls (67%), or emails (19%), with 41% of units adopting all these means of contact.* |
| Chen et al | **Type of service:** General hospital/physical health service<br>**Country:** USA | *The outpatient psychiatry division switched from under 5% virtual visits in March 2019 to over 97% in March 2020. Productivity was maintained at about 95% of previous levels, with 9206 virtual visits in March 2020.*<br>*During the month of March 30 – April 24, 2020, 30% of the virtual visits were conducted via phone.* |
| Childs et al | **Type of service:** CMHT and outpatient services<br>**Country:** USA | *Pre-COVID, 100% of care was delivered in person. The first week after shut-down, telehealth comprised 65.45% of visits (100% over the telephone). In the second week, 91.6% of visits were conducted using telehealth (83.49% over the telephone, 15.6% video). By the third week 99% of appointments used telehealth (30% using video). The percentage of appointments using video increased weekly, peaking at 69.9%* |
| Colle et al | **Type of service:** CMHT and outpatient services<br>**Country:** France | *After 2 weeks of teleconsultations, 376 (91.0%) out of the 413 previously planned appointments were performed.* |
| Connolly et al | **Type of service:** CMHT and outpatient services<br>Veterans Affairs service<br>**Country:** USA | *Daily TMH-V encounters rose from 1,739 on March 11 to 11,406 on April 22 (556% growth, 222,349 total encounters). Between March 11 and April 22, 114,714 patients were seen via TMH-V (Fig. 2). Eighty-eight thousand nine hundred eight (77.5%) were first-time TMH-V users. 12342 MH providers completed a TMH-V appointment between March 11 and April 22 4,281 (34.7%) were first-time TMH-V users. Daily telephone encounters rose from 6,348 on March 11 to 34,396 on April 22 (442% growth).*<br>*Daily in-person encounters fell from 57,296 on March 11 to 10,931 on April 22 (81% decrease).* |
| Dores et al | **Type of service:** Psychology/psychotherapy/<br>counselling service<br>**Country:** Portugal | *During the lockdown period, 17 (15.7%) of the 108 psychologists discontinued therapy and counselling 53 (58.2%) continued to provide services to most or all of their clients. 23 psychologists (25.3%), decreased the number of clients they saw to a range of between 0% and 25%, and for another 15 psychologists (16.5%) that number diminished to a range of between 26% and 50%.* |
| Erekson et al | **Type of service:** Psychology/psychotherapy/counselling service<br>**Country:** USA | Attendance rates for individual therapy temporarily dropped by about 35% but climbed to previous levels within 2 weeks. Group therapy attendance dropped by about 30% but did not fully recover, remaining about 15% lower after 2 weeks. The number of clients receiving individual therapy in 2020 dropped by 43%. During 22 March – 4 April 2020 the service had fewer than half the intakes of any other recent year. |
| Gaddy et al | **Type of service:** Music therapy service<br>**Country:** USA | Of the 869 respondents indicating current contact hours, 70.54% reported that they were providing alternative services, including telehealth services (54.81%), virtual music lessons (17.01%), pre-recorded songs/playlists (16.98%), and pre-recorded video sessions (16.00%). Individual services increased (61.58% (SD = 41.26, whilst group services decreased (24.97% (SD = 37.56)). |
| Gomet et al | **Type of service:** General hospital – addiction service<br>**Country:** France | 100% service users took part in remote care |
| Graell et al | **Type of service:** CMHT and outpatient services<br>Inpatient mental health service<br>**Country:** Spain | During the study period, a total of 1818 outpatient consultations were carried out, 1,329 (73.10%) by telephone or videoconferencing and 489 (26.9%) face-to-face. |
| Grover et al (a) | **Type of service:** CMHT and outpatient services<br>**Country:** India | The majority of the patients reported that they were in touch with their treating doctor (81.5%), with contact initiated by the treating team in 79% of patients. |
| Grover et al (b) | **Type of service:** CMHT and outpatient services<br>Inpatient mental health service<br>Private hospital/clinic<br>**Country:** India | Use of teleservices almost doubled during the lockdown period. 206 (52%) participants provided telecommunication services during the lockdown period. 186 (47%) provided free tele-consultation to the general public and 269 (67.9%) provided free tele-consultation to their patients. 132 (33.3%) were using both voice and video calls (combination of free and paid services). 31 (7.8%) were using only voice calls (combination of free and paid services). |
| Grover et al (c) | **Type of service:** Medical colleges, government-funded institutes mental hospital setting, general hospital psychiatry units<br>**Country:** India | Around 25% of institutes began offering telemental health services. 45.9% of institutes reported that telecommunication services continued during lockdown.<br>Mental health services were being provided to people in quarantine (66.1%) and those with COVID-19 infection (59.6%), family members of patients with COVID19 and those in quarantine (40.4%). |
| Humer et al (a) | **Type of service:**<br>Psychology/psychotherapy/counselling service<br>**Country:** Czech Republic, Germany, Slovakia | Among all countries, the combined (personal contact + telephone + internet) number of patients treated on average per week during COVID-19 ($M = 18.32$, $SD = 12.86$) did not differ from the combined (personal contact + telephone + internet) number of patients treated on average per week in the months before the COVID-19 situation ($M = 19.35$, $SD = 13.73$), $t(337) = -1.506$; $p = 0.133$. |
| Humer et al (b) | **Type of service:**<br>Psychology/psychotherapy/counselling service<br>**Country:** Austria | During the COVID-19 pandemic, face-to-face psychotherapy remained the most abundant treatment modality. |
| Khanna et al | **Type of service:** CMHT and outpatient services<br>**Country:** Australia | There was a 3% increase in appointment bookings compared to the same period in 2019. Cancellation/non-attendance rate dropped from an average of 19% last year to 12% for 2020 |
| Kopec et al | **Type of service:** CMHT and outpatient services<br>**Country:** USA | Prior to COVID-19, Network180 served an average of 2,390 patients/month, which decreased to an average of 1,921 patients/month during the pandemic. This decrease was noted most significantly in crisis services (averaging 822 patients/month before COVID-19 and 640 patients/month during COVID-19).<br>Telehealth increased from 5% of all services prior to COVID-19 to 84% of all services during COVID-19. The majority of services provided via telehealth were audio only (versus audiovisual), with a ratio of 1.9:1 for crisis services and 4:1 for noncrisis services. |
| Looi et al a | **Type of service:** Psychiatrist telehealth service<br>**Country:** Australia | Percentage of consultations conducted using telemental health: ACT: 62% (April) 58% (May) NT: 53% (April) 51% (May) SA: 69% (April) 58% (May) Tasmania: 38% (April) 40% (May) |
| Looi et al b | **Type of service:** Psychiatrist telehealth service<br>**Country:** Australia | The majority of private practice was conducted by telehealth in April but was lower in May as new COVID-19 case rates fell. Percentage of consultations conducted using telemental health NSW: 56% (April) 52% (May) QLD: 63% (April) 53% (May) Vic: 61% (April) 59% (May) WA: |
|  |  | 51% (April) 36% (May) |
| Lynch et al | **Type of service:** CMHT and outpatient services<br>**Country:** USA | *The RS continued providing all services except community-based coaching via telehealth. 90% of CP patients accepted telehealth sessions and maintained their specific treatment plans. Two opted out of telemental health. Mean comparisons between session attendance and cancellations/no-shows during the six-week period before and after telehealth conversion showed no significant differences in service utilization.* |
| Medalia et al | **Type of service:** CMHT and outpatient services<br>**Country:** USA | *Tracking the number of RS enrollees with active participation indicated that in the week before telehealth conversion, when shelter-in-place recommendations commenced, participation dropped from 94% to 52%; after telehealth conversion, participation rose from 67% in the first 4 days to 79% after 1 week and to 84% after 2 weeks.* |
| Miu et al | **Type of service:** CMHT and outpatient services<br>**Country:** USA | *A total of n = 816 participants comprised the analytic sample. A total of n = 400 converted to telehealth and of those n = 64 were SMI. The conversion rates from in-person psychotherapy to teletherapy were similar for SMI (n = 64; 51.6%) and non-SMI (n = 334; 48.3%) groups. The rate at which the SMI group converted from in-person therapy to teletherapy (52%) was not statistically different from that of the non-SMI group (48%) during COVID-19.* |
| Patel et al | **Type of service:** All NHS Trust services<br>**Country:** UK | *From March 2020, in-person contacts reduced substantially from around 9,000 per week to 3,000 per week in early April 2020. Over the same period there was an increase in remote contacts from around 2,500 per week in early March 2020 to around 8,000 per week by the end of April 2020. Total clinical contacts per week dropped from around 12,500 in mid-March to around 10,000 in mid-April 2020. The number of unattended appointments was temporarily reduced in April, May, June and September 2020* |
| Pierce et al | **Type of service:** Variety of MH service settings<br>**Country:** USA | *Psychologists estimated that telepsychology comprised 85.53% of their clinical work during the pandemic, compared with the pre-pandemic context when only 7.07% of their clinical work was conducted remotely.* |
| Probst et al | **Type of service:** Psychology/psychotherapy/counselling service<br>**Country:** Australia | *Face-to-face psychotherapies in personal contact were reduced and remote psychotherapies (via telephone or via Internet) were increased in the early weeks of the COVID-19 lockdown as compared to the months before. Although average increases in psychotherapies via telephone (979%) or via Internet (1561%) were dramatic, there was an undersupply of psychotherapy in Austria in the early weeks of the COVID-19 lockdown as the total number of patients treated on average per week was lower in COVID-19 lockdown than in the months before.* |
| Reilly et al | **Type of service:** Various service types<br>**Country:** USA | *There was uptake of tele-mental health by approximately 80% of respondents by late March or early April 2020. All but 2.11% (19/903) of providers in this study made practice adjustments (transition to tele-mental health).* |
| Rosen et al | **Type of service:** Veterans Health Administration mental health services<br>**Country:** USA | *VHA provided nearly 1.2 million telephone and video encounters to veterans in April 2020 and reduced in-person visits by approximately 80% when compared with the October 2019 to February 2020 period before the pandemic. By June 2020, VHA had an 11-fold increase in encounters using direct-to-home video and a fivefold increase in telephone contacts relative to before the pandemic. VHA reduced in-person visits by approximately 80% when compared with the October 2019 to February 2020 period before the pandemic.* |
| Scharff et al | **Type of service:** Community based training clinic providing therapy<br>**Country:** USA | *The Psychological Services Centre saw an initial retention rate of 82% in the first week of teletherapy, with more clients resuming services in the weeks that followed.* |
| Schlegl et al | **Type of service:** Inpatient mental health service<br>**Country:** Germany | *More than 80% of patients with bulimia nervosa received face-to-face therapy before the COVID-19 pandemic (81.8%) compared to 36.4% during the pandemic (i.e., a decrease by 55.5%). Use of videoconference-based therapy increased from 3.6% to 21.8% and use of telephone contacts from 18.2% to 38.2%, whereas the use of additional online interventions decreased from 3.6% to 0%. Face-to-face psychotherapy decreased by 56% but videoconferencing therapy was only used by 22% of patients.* |
| Sciarrino et al | **Type of service:** Veterans Healthcare Administration<br>**Country:** USA | *Approx. 76% of veterans engaged in PTSD treatment chose to continue despite the COVID-19 pandemic via telehealth.* |
| Sequeira et al | **Type of service:** Residential services | *Post-transition to teletherapy, the average daily virtual program census from 19 March 2020 through 18 April 2020 was 3.3 intensive* |
|  | **Country:** USA | outpatient program patients and 22.4 outpatients. These numbers indicate a slight decrease in IOP patients (−0.3/per day) and an increase in outpatients seen per day (+2.7/per day).<br>There was an increase in the outpatient sessions retained and a decrease in the appointments cancelled. |
| Severe et al | **Type of service:** CMHT and outpatient services<br>**Country:** USA | Take up for remote care was over 95%. 82.8% (n=202) initially chose to receive psychiatric care through video visits, while 13.5% (n=33) chose telephone visits. 1.2% (n=3) decided to postpone care until in-person visit availability. |
| Sharma et al | **Type of service:** General hospital/physical health service<br>**Country:** USA | By March 20, 2020, 67% of all outpatient appointments were conducted at home. Most of these appointments were conducted by phone with some TMH sessions. By March 27, 2020, 90% of all outpatient appointments were done at home, predominantly by phone (59%) but increasingly by HB-TMH (31%). One week later (April 3, 2020), these rates were 48% versus 45%, respectively.<br>By March 31, 2020, 98% of faculty completed expedited training and obtained Departmental approval for HB-TMH services during the COVID-19 crisis. By April 10, 2020, HB-TMH was offered to all established outpatients for individual visits. Only the crisis clinic continued a regular in-clinic presence. |
| Sheehan et al | **Type of service:** CMHT and outpatient services<br>**Country:** UK | 64% were spending at least some time working from their workplace (either solely or in combination with home working). 33.9% were working from home only. Just over a third were at the workplace (n=178, 35.1%) and the remainder (n=147, 28.9%) worked from both home and at the workplace. |
| Termorshuizen et al | **Type of service:** CMHT and outpatient services<br>**Country:** The Netherlands & USA | Most transitioned to telehealth care (USA 45%; Netherlands 42%), with fewer still receiving face-to-face care (USA 3%; Netherlands 6%), or not having been able to engage with their provider at all (USA 6%; Netherlands 5%). |
| Uscher-Pines et al | **Type of service:** CMHT and outpatient services<br>Private hospital/clinic<br>**Country:** USA | Most of the psychiatrists had transitioned to fully virtual practices. Only a quarter of the participants were seeing any patients in person. |
| Uscher-Pines et al | **Type of service:** CMHT and outpatient services<br>Private hospital/clinic; General hospital/physical health service<br>**Country:** USA | Telemedicine use: None (in-person only) 1 (5.6%); Phone only 2 (16.7%); Video only 0 (0.0%); Combination of video and phone 15 (83.3%). |
| van Dijk | **Type of service:** CMHT and outpatient services<br>Psychology/psychotherapy/counselling service<br>**Country:** The Netherlands | Treatment adherence was 100%. |
| Yellowlees et al | **Type of service:** General hospital/physical health service<br>**Country:** USA | By the second day after shutdown, only 8% (N=52) of our appointments were in-person clinic visits, compared with our baseline average of 98%. By the third business day, 100% (N=573) of appointments were conducted virtually, with 92% (N=567) via videoconference and 8% (N=56) by phone. |
| Zulfic et al | **Type of service:** CMHT and outpatient services<br>**Country:** Australia | Some patients still required regular face-to-face reviews, including the 91 patients (29%) who are treated with depot medications and 71 (23%) taking clozapine. |

**Table 5.** Implementation outcomes summary findings for telemental health

| Implementation outcome | Findings | Example studies |
| --- | --- | --- |
| Acceptability | - Remote methods of care are acceptable to most service users and 'exceeded expectations' in terms of satisfaction, but are not viewed as a substitute for face-to-face care.<br>- Clinicians and service users consider the intimacy and connection of face-to-face care is not reproducible on virtual platforms, especially for treatments involving non-verbal communication.<br>- Beyond the pandemic: further data are needed about longer term acceptability, observance, quality of care, and satisfaction.<br>- Clinician burnout due to more appointments per day and requiring more concentration. | (7, 46, 61, 65, 71, 75, 77, 79, 88, 92, 95, 97, 98, 101, 105) |
| Adoption | - Remote working was generally well adopted (most service users switched to remote working).<br>- A few studies also mentioned lower levels of cancellations/DNAs, likely due to not having to travel to the service and the removal of other barriers (e.g. difficulty fitting care around school or work).<br>- Remote working also had the potential to result in reduced waiting times.<br>- Productivity was generally maintained, or in some cases even increased.<br>- Some studies showed no decrease, just change in modality and need to modify psychological treatment. | (7, 51, 53, 55, 57, 59, 62, 76, 80, 82, 83, 114, 115) |
| Appropriateness | - Difficulties managing medication prescription during online consultations.<br>- Concerns around user engagement and assessing new | (57, 61, 79, 83, 100, 101, 113) |
|  | patients.<br>- Harder to assess mental status markers such as hygiene or eye contact, or physical symptoms (e.g. of opioid withdrawal). Though allows to know more about home environment and behaviour outside of clinic.<br>- Does not capture the richness of in-person interaction.<br>- Online felt safer for clinicians providing care to service users at risk for violence and behavioural dysregulation.<br>- Not appropriate for patients with auditory or visual impairments, or with conditions such as migraines. |  |
| Feasibility | - Links with service user and staff needs and resources, in particular problems accessing technology/private space/stable internet connection.<br>- All studies reported good feasibility at least for the short-term emergency response during pandemic.<br>- However, it was not possible to use for specific therapies that require physical presence (role play, collaborative models). Telemental health was less suitable for treating trauma, for clients with severe anxiety, children, and clients with cognitive impairment.<br>- Insurance coverage and legal aspects affected feasibility of implementation in some countries. However, most health insurances caught up and started covering costs. | (7, 57, 59, 63, 65, 74, 75, 79, 83, 95, 97, 105) |
| Fidelity | — No studies explored this area. |  |
| Implementation Cost | - Limited information about cost of intervention, suggested to be 'cost effective' without any presentation of costs.<br>- Reduced travel costs. | (60, 71, 88, 104) |
| Penetration | - Prior to the pandemic, few services used telemental health and for those that did, uptake was low. After the first few weeks, most or all of services were conducted remotely. | (7, 55, 106) |
| Sustainability | - Rates of telemental health use fell as COVID-19 rates declined in the summer of 2020. Links with findings that not all staff and service users would want to continue using remote methods of care after the pandemic ends.<br>- Flexibility is a key advantage of telemental health vs. face-to-face care.<br>- There are some aspects of remote working that services would like to keep, as they provide benefits such as being more efficient and enabling access for certain groups.<br>- Some barriers to remote working (such as lack of experience with online methods of care) have been | (44, 65, 86, 94, 96, 105) |
|  | reduced, making it more likely telemental health will continue to some extent. |  |

Whilst most studies reported high adoption rates, a few studies reported a decrease in attendance: for example, Erekson et al (62) (though possibly because of the university setting) and Dores et al (61) who identified challenges in retention due to: low client adherence, lack of privacy, interruptions at home, lack of appropriate technology and/or simply preference for face-to-face contact.

There was also evidence to indicate that adoption rates of telemental health fell as COVID cases decreased e.g. (85, 86).

#### Appropriateness

There were some concerns raised over the appropriateness of remote care, for example studies reported difficulties managing medication e.g. (46, 109, 110) and concerns around engaging and assessing new patients e.g. (79). Clinicians also found it harder to assess some physical indicators of mental health status (e.g. hygiene, eye contact, physical symptoms of opioid withdrawal), without being able to see the service user in person. However, in contrast, remote methods of working felt safer for clinicians who worked with service users at risk for violence and behavioural dysregulation e.g. (57). Online care was also not necessarily appropriate for patients with auditory or visual impairments, or with conditions such as migraines.

Staff reported concerns around the management of risk and safeguarding of service users when using remote methods of care e.g. (80, 94). Some helpful features of platforms which were thought to improve safety were using the waiting room function, being able to remove call participants, renaming participants (to protect anonymity) and using the private chat function.

#### Feasibility

In general, all studies reported good feasibility, at least for the short-term emergency response during the pandemic. However, some studies reported that telemental health is not suitable for all types of therapy, for example those which require a physical presence (exposure therapy, role play, collaborative models) e.g. (107). Telemental health may also be less suitable for treating trauma (63, 103), for clients with severe anxiety (63), learning difficulties or autism (106), children (63), and clients with cognitive impairment (63, 91).

#### Cost-effectiveness

There was limited information about costs of implementation of remote care in the included studies and no actual costs of telemental health were reported in the papers. However, initial evidence suggests remote care is not a costly intervention, with one paper stating that telemental health is “cost-effective” (70), whilst another mentions the use of “low-cost technologies” used by clinicians (73).

#### Penetration

There was widespread penetration (the extent to which telemental health was integrated into mental health services) of remote methods of care delivery due to the COVID-19 pandemic, despite few services utilising telemental health previously. Services were able to rapidly adapt to this new way of working, with the majority of appointments conducted remotely after the first few weeks of ‘stay at home’ orders.

#### Sustainability

The sustainability of telemental health cannot be completely determined from the included studies, as they present data mostly from the early stages of the COVID-19 pandemic. However, there was some indication that although remote working was widely accepted as a necessity, once restrictions loosened, rates of telemental health use declined (corresponding with the drop in cases in Europe in summer 2020). This correlates with findings that not all staff and service users would want to continue using remote methods of care after the pandemic ends. However, there are some aspects of remote working that both clinicians and service users would like to keep in the future in combination with face-to-face care e.g. (94, 105), as this approach has benefits such as being more efficient, flexible, and enabling access for certain groups e.g. (7, 61, 63).

### Clinical outcomes

Comparing the clinical outcomes of face-to-face and remote care using quantitative measures indicated that telemental health approaches could be as effective as face-to-face care e.g. (56, 104), although it should be noted that most studies were on a small scale. Several studies also reported no psychiatric decompensations after switching to remote care e.g. (87, 89). However, it is important to note that clinical outcomes for telemental health were not comparable for all service users, for example Dores et al (61) found a quarter of psychiatrists reported poorer clinical outcomes after switching to remote care. Another study also indicated that only a third of clinicians felt as though telemental health consultations were comparable to pre-pandemic sessions (115). A full presentation of the clinical outcomes reported in included studies is shown in Table 6.

**Table 6:**
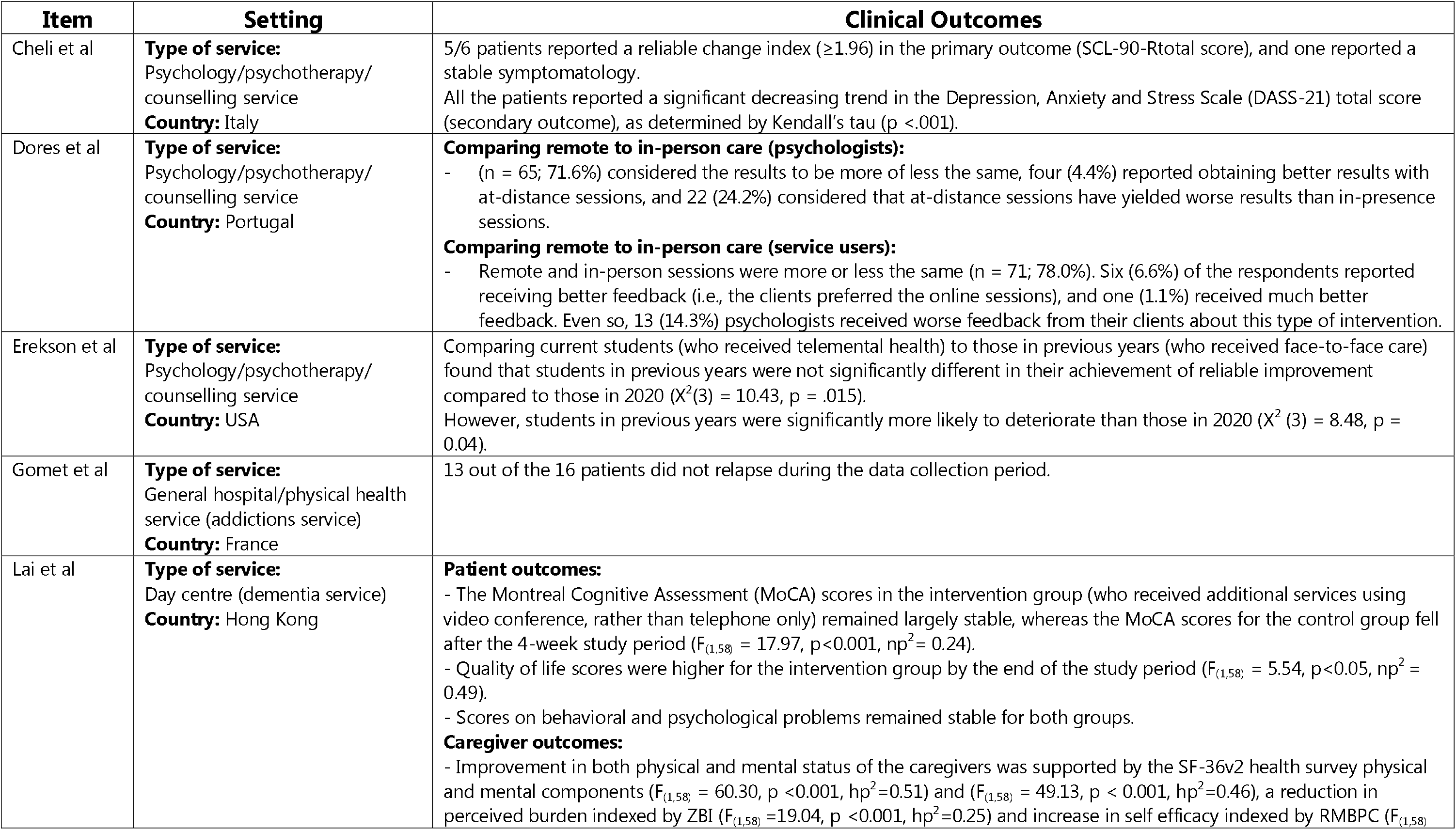
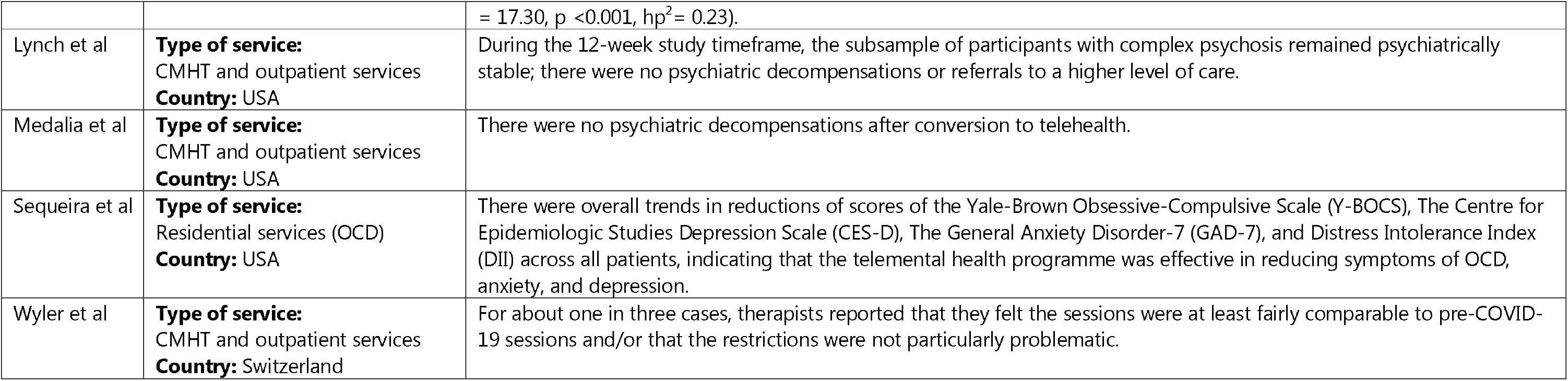
Studies which reported clinical outcomes of telemental health

Although the quality of therapeutic relationships reported by studies was generally good, clinicians reported problems reading patients’ emotions e.g. (44) or feeling less connected to the service user compared to face-to-face sessions e.g. (57). Clinicians also reported difficulties regarding feeling and expressing empathy remotely. Other challenges to therapeutic relationships when using remote care included a lack of client engagement, possible misunderstandings due to lack of non-verbal signals, common context or not having a clear idea of patients’ physical state (alongside reduced privacy).

### Social outcomes

One study (82) compared social outcomes in a trial comparing telephone-only care to caregivers of older adults with neurocognitive disorder, with supplementary video care to both carers and service users. Findings indicated that those who received both telephone and video support had greater resilience, better cognitive functioning and a higher quality of life.

### Organisational and care delivery outcomes

Improved communication was noted between staff when using telemental health when compared to traditional face-to-face care, as the use of online file sharing or discussion platforms facilitated communication between staff e.g. (75, 106). The use of online methods also facilitated staff training and some staff reported that remote working resulted in a better work-life balance e.g. (60).

## Discussion

### Summary of findings

This review collated evidence regarding the implementation and outcomes of remote working in mental health services in the context of the COVID-19 pandemic. Most studies indicate a relatively high level of activity, suggesting that at least in the services studied in higher income countries, mental health care can be shifted to telemental health in a crisis. Services mainly reported using a mixture of phone and video calls, with both service users and clinicians varying in their preference for these modalities. There were some indicators of reduced numbers of missed appointments, potentially due to the greater convenience of remote care, which may make access to services easier for some service users.

There was reasonable acceptability across the studies, at least in conditions where the alternative may have been no contact with services at all. However, there were situations, where telemental health may be less acceptable, including for new patients, physical health aspects of care, and for service users without a private space at home to use for therapy. Telemental health also may not be as feasible for certain types of support, including support which needs a physical presence. There was also evidence that telemental health may not be feasible for some clinical presentations, including some service users with psychosis, learning difficulties or autism. Clinicians also reported a decrease in their ability to develop and maintain a strong therapeutic relationship with service users, due to being unable to pick up on non-verbal cues and a lack of connectedness. The acceptability levels found in this study are not dissimilar to previous studies e.g. (15), even though the participants in the current studies are less likely to be volunteering to pilot a new service and more likely to be using telemental health because they have no alternative.

Few formal investigations of how to improve implementation were identified in this review. However, some strategies for improving adoption/penetration/acceptability may include staff training, the use of telemental health champions, strategies for introducing service users to technology and providing some simple guidance on how to use it best, identifying situations or populations when telemental health is not a good idea and those where it might be better. There was also a lack of fidelity assessments when therapies had to be adapted to fit telemental health delivery formats, therefore little is known about the consequences of these adaptations.

Our interpretation of this pattern of findings is that the successful delivery in a pandemic of telemental health should not necessarily be seen as confirmation that people are happy with this mode of delivery long-term, as some of the identified problems may become more serious over time, and reports of being satisfied may have reflected awareness that at the time of the study, it was difficult to offer care by any other means. The longevity of these changes will ultimately turn not only on information technology, safety, and quality, but also on whether policy changes will support the reimbursements and regulatory adjustments implemented during the current crisis (30, 58).

### Implications for future research

There was a lack of reporting in included studies of trying to identify and reach those patients who are at increased risk of digital exclusion. The needs of those at risk of digital exclusion are still largely underreported in the literature and should be made a priority for future research. Studies also included little information regarding the cost-effectiveness of telemental health implementation. Further research is needed to explore the differences in cost (both to the service and service user) between face-to-face and telemental health care. Further research can also formally compare (rather than simply observe) different delivery support strategies that can improve the implementation and potentially also the clinical effectiveness of telemental health, including for specific conditions and service user groups.

Finally, there is scope to conduct big data studies to identify who is not accessing remote care or those at risk of disengaging, and potential comparisons for matched groups to try to compare effectiveness across a range of settings, as this could be done more quickly than in clinical trials whilst respecting patient preference.

### Strengths and limitations

The studies included in this review identified outcomes across different settings and healthcare systems, which may help findings generalise to different settings. This review also captured recent findings on the use of telemental health during the COVID-19 pandemic, allowing findings to be used to improve both existing and future models of remote mental health care.

However, it is also important to take some limitations into account when interpreting the findings from this review. Firstly, the results from quality assessment indicated that whilst around half of primary research studies and the majority of the service evaluations were high quality, around half of primary research studies were scored as moderate to low. This reflects the short nature of studies and often quick turnaround from data collection to publication. Some studies were also published in preprint form and therefore had not undergone peer review. The majority of studies used cross-sectional data, rather than more rigorous methods. Secondly, there was a lack of high quality quantitative evidence for the clinical effectiveness of telemental health care. Clinical effectiveness outcomes were only reported in 9/77 included studies, with some of these findings only based on qualitative evidence or a small number of service users. It is also important to note that the voices of those who dropped out of care may not be included.

The short time scale for data collection and assessment of changes in practice in included studies could also be viewed as a limitation of this research, as it is not clear if changes will be sustained over time or in other contexts (e.g. lower-income countries). Research was also not inclusive of those not accessing or using remote technologies, meaning there is a risk of those at risk of digital exclusion being forgotten when taking the findings of this review into consideration.

We also designed this review to be conducted rapidly to ensure results would be relevant and quickly available, therefore we chose to search four databases and not all studies were independently double-screened. However, we are confident that the rigour of our searches and inclusion of preprint servers meant that the papers included are representative of the literature on this topic.

## Conclusion

Telemental health was a largely effective method to enable continuation of mental health support during the COVID-19 pandemic. Whilst most reported outcomes were positive, telemental health was not feasible for all types of support and may not be acceptable to all service user groups. A blended approach combining face-to-face and telemental health care may be the most desirable service model for future care. The need remains for higher-quality evidence regarding the clinical effectiveness of telemental health and how uptake can be improved for groups at risk of digital exclusion.

## Data Availability

N/A - systematic review

## Policy Research Unit

This paper presents independent research commissioned and funded by the National Institute for Health Research (NIHR) Policy Research Programme, conducted by the NIHR Policy Research Unit (PRU) in Mental Health. The views expressed are those of the authors and not necessarily those of the NIHR, the Department of Health and Social Care or its arm’s length bodies, or other government departments.

## Applied Research Collaboration South London and King’s Improvement Science

This study is supported by the National Institute for Health Research (NIHR) Applied Research Collaboration South London (NIHR ARC South London) at King’s College Hospital NHS Foundation Trust. NS and LG are members of King’s Improvement Science, which offers co-funding to the NIHR ARC South London and is funded by King’s Health Partners (Guy’s and St Thomas’ NHS Foundation Trust, King’s College Hospital NHS Foundation Trust, King’s College London and South London and Maudsley NHS Foundation Trust), and Guy’s and St Thomas’ Foundation. The views expressed are those of the author(s) and not necessarily those of the NIHR, the NHS, the charities, or the Department of Health and Social Care.

LG is funded by King’s Improvement Science, which is part of the NIHR ARC South London and comprises a specialist team of improvement scientists and senior researchers based at King’s College London. King’s Improvement Science is funded by King’s Health Partners (Guy’s and St Thomas’ NHS Foundation Trust, King’s College Hospital NHS Foundation Trust, King’s College London and South London and Maudsley NHS Foundation Trust), and the Guy’s and St Thomas’ Foundation.

## Conflicts of Interest

NS is the director of the London Safety and Training Solutions Ltd, which offers training in patient safety, implementation solutions and human factors to healthcare organisations and the pharmaceutical industry. The other authors have no conflicts of interest to declare.

## Notes

### Author Declarations

N/A - systematic review

