## Supplementary Table 1 for "Implementation, adoption and perceptions of telemental health during the COVID-19 pandemic: a systematic review"

Supplementary Table 1: Pubmed search strategy

|  | (covid-19[Title/Abstract] OR covid-2019[Title/Abstract] OR covid 2019[Title/Abstract] OR SARS cov 2[Title/Abstract] OR SARS-cov-2[Title/Abstract] OR coronaviru*[Title/Abstract] OR 2019-ncov[Title/Abstract] OR pandemic[Title/Abstract]) |
| --- | --- |
| AND | (mental problem[Title/Abstract] OR mental disorder*[Title/Abstract] OR mental health*[Title/Abstract] OR mental illness[Title/Abstract] OR mentally ill persons[Title/Abstract] OR self-injurious behaviour[Title/Abstract] OR clinical psychology[Title/Abstract] OR mental distress[Title/Abstract] OR mental health service*[Title/Abstract] OR mental health nurs*[Title/Abstract] OR psychiatry[Title/Abstract] OR CAMHS[Title/Abstract] OR psychology[Title/Abstract] OR psychotherap*[Title/Abstract] OR dementia[Title/Abstract] OR Alzheimer*[Title/Abstract] OR depression[Title/Abstract] OR depressive disorder[Title/Abstract] OR anxiety[Title/Abstract] OR anxiety disorder[Title/Abstract] OR phobi*[Title/Abstract] OR agoraphobi*[Title/Abstract] OR anxious[Title/Abstract] OR obsess*[Title/Abstract] OR compulsi*[Title/Abstract] OR panic[Title/Abstract] OR PTSD[Title/Abstract] OR post traumatic stress[Title/Abstract] OR posttraumatic stress[Title/Abstract] OR stress disorder*[Title/Abstract] OR psychiatr*[Title/Abstract] OR SMI[Title/Abstract] OR psycho*[Title/Abstract] OR schizo*[Title/Abstract] OR manic[Title/Abstract] OR mania[Title/Abstract] OR bipolar[Title/Abstract] OR personality disorder*[Title/Abstract] OR self-harm[Title/Abstract] OR self-injury[Title/Abstract] OR psychological disorder[Title/Abstract] OR psychiatric illness[Title/Abstract] OR psychiatric disorder[Title/Abstract] OR bipolar disorder[Title/Abstract] OR eating disorder[Title/Abstract] OR neurotic disorder[Title/Abstract] OR psychotic disorder[Title/Abstract]) OR (dementia[MeSH Subheading]) OR (Alzheimer disease[MeSH Subheading])) OR (anxiety disorders[MeSH Subheading]) OR (bipolar disorder[MeSH Subheading]) OR (feeding[MeSH Subheading] AND eating disorders[MeSH Subheading]) OR (depressive disorder[MeSH Subheading]) OR (neurotic disorders[MeSH Subheading]) OR (personality disorders[MeSH Subheading])) OR (psychotic disorders[MeSH Subheading]) OR (schizophrenia[MeSH Subheading]) OR (mental disorders[MeSH Subheading]) OR (mental health[MeSH Subheading]) OR (mentally ill persons[MeSH Subheading]) OR (self-injurious behaviour[MeSH Subheading]) OR (psychology, clinical[MeSH Subheading]) OR (mental health services[MeSH Subheading]) OR ("dementia"[MeSH Terms]) OR ("Alzheimer disease"[MeSH Terms]) OR ("anxiety disorders"[MeSH Terms]) OR ("bipolar disorder"[MeSH Terms])) OR ("feeding and eating disorders"[MeSH Terms]) OR ("depressive disorder"[MeSH Terms]) OR ("neurotic disorders"[MeSH Terms]) OR ("personality disorders"[MeSH Terms]) OR ("psychotic disorders"[MeSH Terms]) OR ("schizophrenia"[MeSH Terms]) OR ("mental disorders"[MeSH Terms])) OR ("mental health"[MeSH Terms])) OR ("mentally ill persons"[MeSH Terms]) OR ("self-injurious behaviour"[MeSH Terms]) OR ("psychology, clinical"[MeSH Terms]) OR ("mental health services"[MeSH Terms]) |
| AND | (telemedicine[Title/Abstract] OR remote consultation[Title/Abstract] OR distance counselling[Title/Abstract] OR computer-assisted therapy[Title/Abstract] OR videoconferenc*[Title/Abstract] OR video-conferenc*[Title/Abstract] OR video call[Title/Abstract] OR video-call[Title/Abstract] OR video-based call[Title/Abstract] OR "video based call"[Title/Abstract] OR videophone[Title/Abstract] OR video phone[Title/Abstract] OR internet-based intervention[Title/Abstract] OR emedicine[Title/Abstract] OR e-medicine[Title/Abstract] OR etherap*[Title/Abstract] OR e-therap*[Title/Abstract] OR telepsychiatry[Title/Abstract] OR telecare[Title/Abstract] OR telecommunication[Title/Abstract] OR teleconferencing[Title/Abstract] OR teletherap*[Title/Abstract] OR tele-therap*[Title/Abstract] OR telemental[Title/Abstract] OR tele-mental[Title/Abstract] OR online care [Title/Abstract] OR online therapy [Title/Abstract] OR online treatment [Title/Abstract] OR remote care[Title/Abstract] OR email[Title/Abstract] OR e-mail[Title/Abstract] OR consult*[Title/Abstract] OR counsel*[Title/Abstract] OR CBT[Title/Abstract]) ("Telemedicine"[MeSH Terms]) OR ("remote consultation"[MeSH Terms]) OR ("distance counselling"[MeSH Terms]) OR ("therapy, computer assisted"[MeSH Terms]) OR ("videoconferencing"[MeSH Terms]) OR ("internet-based intervention"[MeSH Terms]) |
| AND | Filters: from 2020/1/1 - 2020/12/8 |
