## Supplementary Table 2 for "Implementation, adoption and perceptions of telemental health during the COVID-19 pandemic: a systematic review"

|  | **Study** | **Report Type** | **Aim of study** | **Modality used** | **Country/countries involved** | **Nature of mental health problem/diagnosis & Type of service** | **Characteristics of participants and sample size** |
| --- | --- | --- | --- | --- | --- | --- | --- |
| 1 | Aafjes-van Doorn et al (2020) | Primary research study | Survey of therapists’ experiences of video therapy during the pandemic. | Video | USA, Canada Europe: Hungary, Italy, UK, Germany, Norway, Sweden, Switzerland, Latvia, Ireland | Not stated  Psychology/psychotherapy/  counselling service | Staff  Counsellors, psychologists  female (N = 105; 74.5%)  White (N = 120; 85.7%). Age range 23-79, average age 46 (SD = 14.83).  Sample size: 144 |
| 2 | Anton et al, (2020) | Service evaluation/audit | Description of transition to telemedicine | Video  Phone  Text messaging  Email  Mixed face-to-face and telehealth | USA | Depression, PTSD  General hospital/physical health service | Staff  Physicians, nurses, social workers, psychologists |
| 3 | Barney et al (2020) | Service evaluation/audit | Description of transition to telemedicine | Video | USA | Mixed secondary mental health service users  CMHTs and outpatient services | Staff  Nurses, doctors, medical assistants, front desk staff |
| 4 | Bekes et al ^a^ (2020) | Primary research study | Survey of psychotherapists’ attitudes toward online psychotherapy | Video | Canada, USA, Europe (countries not stated) | Mixed primary care or psychology/psychotherapy service users  Psychology/psychotherapy/counselling service Private hospital/clinic CMHTs and outpatient services | Staff  Psychotherapists Mean age 46.50 years (SD 14.83, range 23–79), female (106; 75.2%)  White (120; 82.8%)  Sample size: 145 |
| 5 | Békés et al ^b^ (2020) | Primary research study | Survey of psychoanalytical therapists' experiences of videoconference therapy during the pandemic | Video  Phone | Canada, USA, Europe (countries not stated) | Mixed primary care or psychology/psychotherapy service users  CMHTs and outpatient services Psychology/psychotherapy/counselling service Private hospital/clinic | Staff  Psychologists, medical doctors, social workers, counsellors, family therapists, other psychoanalyst/therapists  Female 66.8% Male 33.2%  Ethnicity White 93.4%, Hispanic or Latino 3.7%, Asian or Asian Indian 3.2%, Middle Eastern 1.6%, American Indian or Alaska Native 1.1%, African American 1.1%  Average age 60.5 (SD = 15; range, 28–90)  Sample size: 190 |
| 6 | Benaque et al, (2020) | Service evaluation/audit | Description of service changes due to the pandemic | Video  Phone  Text messaging | Spain | Dementia  Voluntary sector/non-profit organisation. | Staff  Neurologists, geriatricians, neuropsychologists, social workers, nurses, and clinical and technical staff |
| 7 | Berdullas Saunders et al (2020) | Service evaluation/audit | Description of the use of a psychological helpline | Phone | Spain | General population experiencing: depression, anxiety, OCD, self harm. Worsening pre-existing mental health problems.  Helpline | General population  15170 calls, of which 11417 resulted in psychological interventions.  Users included the general population, healthcare and other essential professionals and the patients and relatives of the sick or deceased.  Female 73.5% Male 26.5%  Average age: 49 (SD =14) |
| 8 | Bhome et al (2020) | Primary research study | Survey of staff perspectives on delivery of services to older adults during the pandemic | Not applicable | UK | Dementia  CMHTs and outpatient services. Inpatient mental health service | Staff  Psychiatrists, nurses, occupational therapists, peer workers, psychologists, social workers, managerial or lead clinician role Gender: 81% female, ethnicity: 93% White  Sample size: 158 |
| 9 | Bierbooms et al, (2020) | Primary research study | Interviews with health professionals on the sustainability of online treatment after the pandemic | Video  Text messaging Online treatment modules | The Netherlands | General population  CMHTs and outpatient services | Staff  Health professionals of all types  Sample size: 11 |
| 10 | Boldrini et al (2020) | Primary research study | Survey of psychotherapists' experience with telepsychotherapy during the pandemic | Video  Phone  Mixed face-to-face and telehealth | Italy | Mixed primary care or psychology/psychotherapy service users  CMHTs and outpatient services Psychology/psychotherapy/counselling service Private hospital/clinic | Staff  Psychotherapist or analyst  84% female; mean age = 45.1 (SD = 10.2)  Sample size: 308 |
| 11 | Burton et al (2020) | Primary research study | Interviews with people with mental health conditions on their experience during the pandemic | Not applicable | UK | Mixed secondary mental health service users  No specific setting | Service users  Mean age 43, range 23-70, female 59.1%, White: 68.2%, Asian (Indian/Pakistani) 13.6%, Mixed White/Asian9.1%, Black British/Black African 9.1%  Sample size: 22 |
| 12 | Carpiniello et al (2020) | Letter (with data) | Survey to explore the impact of the pandemic on the functioning of Mental health services | Not applicable | Italy | Mixed secondary mental health service users  CMHTs and outpatient services Inpatient mental health service Residential services | Staff  Heads of Mental Health departments  Sample size: 71 |
| 13 | Cheli et al (2020) | Primary research study | Evaluation of a crisis intervention for patients diagnosed with psychosis | Video | Italy | Psychosis and bipolar  Psychology/psychotherapy/  counselling service | Service users  Male to female ratio 4:2, age range 19 – 27  Sample size: 6 |
| 14 | Chen et al (2020) | Commentary/editorial (with data) | Description of changes made to mental health services due to the pandemic | Video  Phone | USA | Mixed secondary mental health service users  General hospital/physical health service | Not applicable – description of service change |
| 15 | Childs et al (2020) | Service evaluation/audit | Description of changes made in an outpatient psychiatric service due to the pandemic | Video  Phone | USA | Mixed secondary mental health service users  CMHTs and outpatient services | Service users  Age range 13-17 |
| 16 | Colle et al (2020) | Letter (with data) | Evaluation of teleconsultation during the pandemic | Video  Phone | France | Mixed secondary mental health service users  CMHTs and outpatient services | Service users  Mean age 46, Female 57.1% |
| 17 | Connolly et al (2020) | Service evaluation/audit | Description of changes to services during the pandemic | Video  Phone | USA | Mixed secondary mental health service users  CMHTs and outpatient services Veterans Affairs service | Not applicable – description of service change |
| 18 | Datta et al (2020) | Service evaluation/audit | Description of transition to telehealth during the pandemic | Video | USA | Eating disorders  Inpatient mental health service | Not applicable – description of service change |
| 19 | Dores et al (2020) | Primary research study | Exploration of mental health professionals’ attitudes regarding ICT use | Video  Phone  Social networks  Email | Portugal | Mixed secondary mental health service users  Psychology/psychotherapy/  counselling service | Staff  Psychologists  Female (89, 82.4%). mean age 37.20 years old (SD = 10.05; range 23-65, Sample size: 108 |
| 20 | Erekson et al (2020) | Service evaluation/audit | Exploration of use of telehealth in a student counselling service during the pandemic | Video | USA | Student counselling services  Psychology/psychotherapy/  counselling service | Staff  Therapists |
| 21 | Feijt et al (2020) | Primary research study | Exploration of staff experiences of online treatment during the pandemic | Video  Phone  Chat sessions  Email | The Netherlands | Mixed secondary mental health service users  Not stated/unclear | Staff  Healthcare professionals  Female 82 %, average age 38, range 25- 60  Sample size: 51 |
| 22 | Fernandez et al (2020) | Primary research study | Survey on the impact of the pandemic for people diagnosed with an eating disorder | Not stated | Spain | Eating disorders  Inpatient mental health services | Service users  Mean age 33.4, SD 15.8, range: 13-77, Female 86% Sample size: 121 |
| 23 | Foye et al (2020) | Primary research study | Exploration of impact of the pandemic on mental health nurses | Video  Phone | UK | Mixed secondary mental health service users  CMHTs and outpatient services Crisis and emergency mental health services Inpatient mental health service | Staff  Nurses  Gender: 77.9% female  Age: 3.1% <25; 19% 25-34; 24.1% 35-44; 35.2% 45-54; 18.2% 55-64; 0.4% >65  Ethnicity: 88.5% White; 1.7% Asian; 5.7% Black; 2.9% Mixed;1.1% other/prefer not to say |
| 24 | Gaddy et al (2020) | Primary research study | Exploration of the impact of the pandemic on music therapy professionals | Video  Phone | USA | Not stated  Music therapy | Staff  Staff group: Music therapists  91.01% female, 8.31% male, 0.68% transgender non-conforming.  Age range 18-84  Sample size: 1196 |
| 25 | Gillard et al (2020) | Primary research study | Exploration of the experiences of people with mental health problems during the COVID-19 pandemic. | Video  Phone  Text messaging | UK | Mixed secondary mental health service users  CMHTs and outpatient services Primary care Inpatient mental health service Voluntary sector/non-profit organisation Psychology/psychotherapy/counselling service | Service users  18-24 4%; 25-34 31%; 35-44 24%; 45-54, 22%; 55-69 6%; >70 8%  White British 53%; White Other 10%; Mixed/Multiple ethnic groups 8%; Asian/Asian British 12%; Black / Black British 14%; Other ethnic group 2%. -Female 69% |
| 26 | Gomet et al (2020) | Service evaluation/audit | Description and review of the implementation of remote working in an addiction outpatient service | Phone | France | Substance abuse  General hospital/physical health service | Service users |
| 27 | Graell et al (2020) | Service evaluation/audit | Exploration of the impact of the pandemic on a Child and Adolescent Eating Disorders service | Video  Phone  Mixed face-to-face and telehealth | Spain | Child and Adolescent Eating disorders  CMHTs and outpatient services Inpatient mental health service | Service users  Day hospital: Mean age 13.18, female 92.9%; Outpatients: mean age 14.74, female 87.3%  Sample size: Day hospital: 27, Outpatients: 338 |
| 28 | Grover et al ^a^ (2020) | Letter (with data) | Evaluation of the monitoring of patients with schizophrenia on clozapine during the pandemic | Phone  Text messaging | India | Psychosis and bipolar  CMHTs and outpatient services | Service users  Mean age 33.7, Male 55.1% |
| 29 | Grover et al ^b^ (2020) | Primary research study | Evaluation of the impact of the pandemic on mental health services in India | Video  Phone | India | Mixed secondary mental health service users  CMHTs and outpatient services Inpatient mental health service Private hospital/clinic | Staff  Psychiatrists  Sample size: 396 |
| 30 | Grover et al ^c^ (2020) | Primary research study | Evaluation the impact of the pandemic on mental health services in India | Video  Phone | India | Mixed secondary mental health service users  Medical colleges, government-funded institutes mental hospital setting, general hospital psychiatry units | Staff  Professors and Heads of Department  Sample size: 109 |
| 31 | Haxhihamza et al (2020) | Service evaluation/audit | Evaluation of the satisfaction of patients with telepsychiatry due to the pandemic. | Mixed face-to-face and telehealth | North Macedonia | Mixed secondary mental health service users  Day hospital | Service users  Mean age 40.25, male n=18, female=11  Sample size: 28 (inconsistency in how this was reported in the paper) |
| 32 | He et al (2020) | Primary research study | Evaluation of a psychological intervention programme | Video  Phone  Mixed face-to-face and telehealth  Social media | China | General population  No specific setting/general population  Helplines  Online media programs | Not stated |
| 33 | Hom et al (2020) | Service evaluation/audit | Description of the development of a virtual Program for an acute psychiatric population | Video | USA | Mixed secondary mental health service users  Private hospital/clinic | Staff  Psychologists, psychiatrists, nurses, counsellors  Service users  Mean age 39.04, range 19-69; female 52% |
| 34 | Humer et al (2020) | Primary research study | Survey of psychotherapists’ views on working during the pandemic | Video  Phone  Email  Mixed face-to-face and telehealth | Czech Republic Germany Slovakia | Not stated  Psychology/psychotherapy/counselling service | Staff  Psychotherapist or analyst  Mean age 46.70 (SD = 10.68), female 77.8% Sample size: 338 |
| 35 | Humer et al (2020) | Primary research study | Survey of psychotherapists view on the use of the internet during the pandemic | Video | Austria | Not stated  Psychology/psychotherapy/counselling service | Staff  Psychotherapist or analyst  Average age 51.7, female 75.7%  Sample size: 1547 |
| 36 | Izakova et al (2020) | Primary research study | Survey of the impact of the pandemic on mental health experts | Video  Phone | Slovakia | Not stated  CMHTs and outpatient services  Inpatient mental health service | Staff  Psychiatrists, psychotherapist or analyst, psychologist  female 79%  Sample size: 157 |
| 37 | Johnson et al (2020) | Primary research study | Survey of the experiences of mental health staff during the pandemic | Video  Phone  Mixed face-to-face and telehealth | UK | Mixed secondary mental health service users  Residential services Voluntary sector/non-profit organisation CMHTs and outpatient services  Crisis and emergency mental health services Inpatient mental health service Psychology/psychotherapy/counselling service | Staff  Peer workers, psychologists, social workers, psychiatrists, nurses, managers  female 80.0% white ethnic groups 87.0%  Sample size: 2180 |
| 38 | Jurcik et al (2020) | Primary research study | Exploration of how the pandemic affected mental health services | Video  Phone  Mixed face-to-face and telehealth | Australia Canada Japan Russia | Mixed secondary mental health service users  Not stated/unclear | Staff  Psychologists and psychologists  Female 62.5%, mean age: 41.75; SD = 7.61  Sample size: 8 |
| 39 | Khanna et al (2020) | Service evaluation/audit | Description of services changes in a trauma service during the pandemic | Video  Phone  Mixed face-to-face and telehealth | Australia | PTSD  CMHTs and outpatient services | Staff  Psychiatrists and psychologists  Sample size: 21 |
| 40 | Kopec et al (2020) | Service evaluation/audit | Description of the transition to telehealth in a community mental health service | Video  Phone | USA | Mixed secondary mental health service users  CMHTs and outpatient services | Not applicable – description of service change |
| 41 | Lai et al (2020) | Primary research study | Evaluation of the benefits of telehealth to people with Dementia and their carers | Video  Phone | Hong Kong | Dementia  Day centre | Service users  Age range 64 -80, female:male ratio 18:12 Carers  Age range 66-82, female:male ratio 18:12  Sample size: 60 |
| 42 | Lakeman & Crighton (2020) | Primary research study | Exploration of providing Dialectical Behaviour Therapy using telehealth technology | Video  Phone  Mixed face-to-face and telehealth | Australia | Personality disorder  Psychology/psychotherapy/  counselling service | Staff  Psychologists, social workers, nurses, occupational therapists  Average age 47; range 25–64 years  Sample size: 28 |
| 43 | Lin et al (2020) | Primary research study | Evaluation of psychological hotline services set up during the pandemic | Phone  Text messaging | China | General population  Helplines | Not applicable – evaluation of a new service |
| 44 | Looi et al^a^ (2020) | Primary research study | Evaluation of the use of psychiatry telehealth in smaller states | Video  Phone  Mixed face-to-face and telehealth | Australia | Mixed secondary mental health service users  Psychiatrist telehealth service | Staff  Psychiatrists |
| 45 | Looi et al^b^ (2020) | Primary research study | Evaluation of the use of psychiatry telehealth in larger states | Video  Phone  Mixed face-to-face and telehealth | Australia | Mixed secondary mental health service users  Psychiatrist telehealth service | Staff  Psychiatrists |
| 46 | Lynch et al (2020) | Service evaluation/audit | Description of change to telehealth in a service for people with psychosis | Video | USA | Psychosis and bipolar  CMHTs and outpatient services | Service users  Female n=22, Male n=37, Transgender/non-binary/other n=5  White n=59, Black/African American n=1, Hispanic n=2, Asian n=2  Sample size: 64 |
| 47 | McBeath et al (2020) | Primary research study | Exploration of the experiences of psychotherapists working remotely during the pandemic | Video  Phone  Text messaging  Email | UK | Mixed psychology service users  Psychology/psychotherapy/  counselling service | Staff  Psychotherapists  Characteristics  female 79%, male 18%, ‘other’ 3%  Sample size: 335 |
| 48 | Medalia et al (2020) | Commentary/editorial (with data) | Description of the change to telehealth in a service for people with serious mental illness | Video | USA | Mixed secondary mental health service users  CMHTs and outpatient services | Not applicable—description of service change |
| 49 | Miu et al (2020) | Service evaluation/audit | Evaluation of the engagement with telehealth of people with SMI during the pandemic | Video  Phone | USA | Mixed secondary mental health service users  CMHTs and outpatient services | Staff  Counsellors, psychologists, social workers  Female 75%, White 79%  Sample size: 24 |
| 50 | Olwill et al (2020) | Primary research study | Survey of psychiatrists’ experience of remote consultations | Phone | Ireland | Mixed secondary mental health service users  CMHTs and outpatient services | Staff  Psychiatrists  Age 25–34: 38%, 35–44: 35% ,45–54: 8% 55–64, 15%  Sample size: 26 |
| 51 | Patel et al (2020) | Primary research study | Analysis of Health Record data on the impact of remote consultation during the pandemic | Not applicable | UK | Mixed secondary mental health service users  All NHS Trust services | Not applicable – description of whole service health record date |
| 52 | Peralta et al (2020) | Primary research study | Evaluation of the effectiveness of teleconsultation use during the pandemic | Video  Phone  Text | Dominican Republic | General population  Voluntary sector/non-profit organisation Helplines | Not stated  6800 interventions  67.3% of the interventions were requested by women. 77.8% between 18 and 59. |
| 53 | Pierce et al (2020) | Primary research study | Survey of the impact of telepsychology use by psychologists before and during the pandemic | Video  Phone | USA | Mixed primary care or psychology/psychotherapy service users  General hospital/physical health service CMHTs and outpatient services Residential services Psychology/psychotherapy/counselling service | Staff  Psychologists  Age =57.29 (SD=11.42); Men: 35.4% Women: 64.2% Genderqueer: 0.1% Gender nonconforming: 0.1% Transman: 1.0%  Race/ethnicity: White/European-American 91.1%, Latinx/Hispanic 2.3%, Asian/Asian-American 1.9%, Black/African-American 1.9%, Multiracial/multiethnic 1.5%, Other 1.1%, American Indian/Alaska Native/Native American 0.1%,  Sample size: 2619 |
| 54 | Probst et al (2020) | Primary research study | Investigation of changes to psychotherapy compared to the months before the pandemic | Phone | Austria | Mixed primary care or psychology/psychotherapy service users  Psychology/psychotherapy/counselling service | Staff  Psychotherapists  Mean age 51.6, SD 9.69 75.7% female  Sample size: 1547 |
| 55 | Reilly et al (2020) | Primary research study | Survey to understand change in practice by healthcare staff during the pandemic | Not stated | USA | Mixed primary care and  secondary mental health service users  CMHTs and outpatient services Primary care Private hospital/clinic General hospital/physical health service Psychology/psychotherapy/counselling service  Other (specify) Veterans hospital or military hospital/clinic Prison | Staff  Psychologist, social workers, counsellors, peer workers, psychiatrists, nurses  Mean age 39.50 SD 11.50  Male 16.50%, female 82.95% Transgender man 0.22% Genderqueer/nonconforming 0.33%  American Indian/Alaska Native 0.11% Asian/Asian American 3.22% Black/African American 3.22% Hispanic/Latinx 3.67% White 86.78% Multiracial 2.78% Different racial identity (ie, Arab, Jewish, Mestiza) 0.22%  Sample size: 903 |
| 56 | Roach et al (2020) | Primary research study | Interviews to understand the experience of people with dementia during the pandemic | Not applicable | Canada | Dementia  CMHTs and outpatient services | Service users  Mean age 69, SD 8.3, Female 50%  Sample size: 21 |
| 57 | Roncero et al (2020) | Service evaluation/audit | Description of the response of a mental health network to the pandemic. | Video  Phone  Mixed face-to-face and telehealth | Spain | Mixed primary care and  secondary mental health service users  CMHTs and outpatient services Crisis and emergency mental health services General hospital/physical health service Inpatient mental health service | Not applicable – description of service change |
| 58 | Rosen et al (2020) | Service evaluation/audit | Description of transition to telemental health services | Video  Phone | USA | Mixed primary care and  secondary mental health service users  Veterans Health Administration mental health services | Not applicable – description of service change |
| 59 | Sasangohar et al (2020) | Service evaluation/audit | Description of implementation of telepsychiatry in a psychiatric practice | Video  Phone  Email | USA | Mixed secondary mental health service users  CMHTs and outpatient services | Not applicable – description of service change |
| 60 | Scharff et al (2020) | Service evaluation/audit | Description of changes made by a Psychological Service during the pandemic | Video | USA | Mixed primary care and  secondary mental health service users  community based training clinic providing therapy | Not stated |
| 61 | Schlegl et al (2020) | Primary research study | Survey to investigate the impact of the pandemic on patients with Bulimia Nervosa | Not applicable | Germany | Eating disorders  Inpatient mental health service | Service users  Mean age 24.42, SD 6.36, range 17-46  100% female  Sample size: 55 |
| 62 | Sciarrino et al (2020) | Commentary/editorial (with data) | Description of providing trauma-focused treatment using telehealth during the pandemic. | Video  Apps | USA | PTSD  General hospital/physical health service Veterans Healthcare Administration | Not stated |
| 63 | Sequeira et al (2020) | Service evaluation/audit | Description of change to services for people with OCD during the pandemic | Video | USA | OCD  Residential services | Service users  Age range 17-34  80% White, 20% biracial , Female n=2, male n=3  Sample size: 5 |
| 64 | Severe et al (2020) | Primary research study | Survey of patients using a mental health service to explore decisions to accept or decline telepsychiatry | Video  Phone | USA | Mixed secondary mental health service users  CMHTs and outpatient services | Service users  Age from under 12 – 85, 77.5% White, 10.7% Black/African American, 4.5% Asian, female 68.4%  Sample size: 244 |
| 65 | Sharma et al (2020) | Service evaluation/audit | Description of the implementation of a home-based telemental health service during the pandemic | Video  Phone | USA | Child and adolescent services  General hospital/physical health service | Staff  Psychologists, psychiatrists, analysts, nurses  Sample size: 105 |
| 66 | Sheehan et al (2020) | Primary research study | Survey of the experiences of staff working with people with intellectual and other developmental disabilities | Not applicable | UK | Intellectual disabilities  CMHTs and outpatient services | Staff  Nurses, psychologist, social workers, support workers, psychiatrists  Female 78.9%, Aged 25 – 64, White ethnicity 82.2% Sample size: 648 |
| 67 | Sklar et al | Primary research study | Exploring the impact of the pandemic on mental health services in Indiana | Not applicable | USA | Not stated  CMHTs and outpatient services | Staff |
| 68 | Termorshuizen et al (2020) | Primary research study | Survey to evaluate the impact of the pandemic on people with eating disorders | Not applicable | The Netherlands USA | Eating disorders  CMHTs and outpatient services | Service users  Age range 16 to over 60, female USA 97%, Netherlands 99%  Sample size: USA 511, Netherlands 510 |
| 69 | Uscher-Pines et al^a^ (2020) | Primary research study | Interviews with psychiatrists to understand how change in delivery has affected mental health care | Video  Phone | USA | Mixed secondary mental health service users  CMHTs and outpatient services Private hospital/clinic | Staff  Psychiatrists  Sample size: 20 |
| 70 | Uscher-Pines et al^b^ (2020) | Primary research study | Interviews with clinicians to understand the experience of using telemedicine for Opiate Use Disorder | Video  Phone | USA | Opiate Use Disorder  CMHTs and outpatient services Private hospital/clinic General hospital/physical health service | Staff  Physicians, psychiatrists  Sample size: 18 |
| 71 | van Dijk et al (2020) | Service evaluation/audit | Description of transforming a day-treatment program for older people into an online program | Video | The Netherlands | Mixed secondary mental health service users  CMHTs and outpatient services  Psychology/psychotherapy/counselling service | Staff  Psychologists, nurse practitioners, art therapists, psychomotor therapists |
| 72 | Wang et al (2020) | Primary research study | Survey to compare Chinese and US practitioners' attitudes towards teletherapy during the pandemic | Video  Phone | USA China | Mixed primary care or psychology/psychotherapy service users  Not stated/unclear | Staff  Counsellors, psychoanalytic practitioners, psychologists, social workers, psychiatrists  Age 20‐29yrs USA.64% China 66% 30-39yrs USA 52% China 52% 40-49yrs USA 34.55% China 35% 50-59yrs USA 9.7% China 9.7%  Gender Female USA 71.52% China 73.17%  Sample size: 329 |
| 73 | Wilson et al (2020) | Primary research study | Survey to explore staff perceptions of the impact of the pandemic on perinatal services | Video  Phone  Mixed face-to-face and telehealth | UK | Perinatal services  CMHTs and outpatient services Crisis and emergency mental health services Inpatient mental health service | Staff  Psychologists, social workers, psychiatrists, nurses, occupational therapists  85.2% female, 70.3% White British.  Sample size: 363 |
| 74 | Wood et al (2020) | Service evaluation/audit | Description of the implementation of group teletherapy for people with first episode psychosis | Video | USA | Psychosis and bipolar  CMHTs and outpatient services | Service users  Mean age26.9, SD 4.8, range 18-29, 55% Male  Sample size: 7 |
| 75 | Wyler et al (2020) | Primary research study | Exploration of the experience of therapy sessions for people with ADHD and their therapists during the pandemic | Video  Phone | Switzerland | ADHD  CMHTs and outpatient services | Staff  Therapists  Service users  Mean age 39.10, SD 11.19, 55% Male  Sample size: 60 therapist/service user dyads |
| 76 | Yellowlees et al (2020) | Commentary/editorial (with data) | Description of the rapid conversion of an outpatient psychiatric clinic to a telepsychiatry clinic | Video  Phone | USA | Mixed secondary mental health service users  General hospital/physical health service | Not applicable – description of service change |
| 77 | Zulfic et al (2020) | Letter (with data) | Audit to understand the move to telephone support for people using a Community Mental Health Team | Phone | Australia | Mixed secondary mental health service users  CMHTs and outpatient services | Service users  Sample size: 314 |
